# Tracking and predicting the African COVID-19 pandemic

**DOI:** 10.1101/2020.11.13.20231241

**Authors:** Paddy Ssentongo, Claudio Fronterre, Andrew Geronimo, Steven J. Greybush, Pamela K. Mbabazi, Joseph Muvawala, Sarah B. Nahalamba, Philip O. Omadi, Bernard T. Opar, Shamim A. Sinnar, Yan Wang, Andrew J. Whalen, Leonhard Held, Chris Jewell, Abraham J. B. Muwanguzi, Helen Greatrex, Michael M. Norton, Peter Diggle, Steven J. Schiff

## Abstract

The ongoing coronavirus disease 2019 (COVID-19) pandemic is heterogeneous throughout Africa and threatening millions of lives. Surveillance and short-term modeling forecasts are critical to provide timely information for decisions on control strategies. We use a model that explains the evolution of the COVID-19 pandemic over time in the entire African continent, parameterized by socioeconomic and geoeconomic variations and the lagged effects of social policy and meteorological history. We observed the effect of the human development index, containment policies, testing capacity, specific humidity, temperature and landlocked status of countries on the local within-country and external between-country transmission. One week forecasts of case numbers from the model were driven by the quality of the reported data. Seeking equitable behavioral and social interventions, balanced with coordinated country-specific strategies in infection suppression, should be a continental priority to control the COVID-19 pandemic in Africa.

## 1 Introduction

The ongoing coronavirus disease 2019 (COVID-19) pandemic in Africa is threatening millions of lives, a crisis compounded by the continent’s unique spectrum of disease and fragile health care infrastructure (*1*). Essential to African countries’ efforts to control the pandemic are effective methods to track and predict new cases and their sources in real time. Time-critical interpretation of daily case data is required to inform public health policy on mitigation strategies and resource allocation. To address this need, we developed a data-driven disease surveillance framework to track and predict country-level case incidence from internal and external sources. We chose a spatiotemporal strategy to take advantage of and combine openly available data on coronavirus epidemiology, social policy affecting human movement and public health, meteorological factors, socioeconomic and demographic variables, seeking to inform rapid policy development.

The first COVID-19 case on the continent was reported in Egypt on February 14, 2020. By August 13, 2020, over 1 million new cases and over 20,000 deaths had been reported in all African Union (AU) Member States according to the Africa Centres for Disease Control and Prevention (CDC) (https://africacdc.org/covid-19/). Over 44 million cases and 190,000 deaths in Africa are projected within the first year of the pandemic (*2*). Although Africa has a younger age distribution that could theoretically lead to fewer symptomatic or severe infections (*3*), modeling predicts that the relatively low healthcare capacity in many parts of Africa, in combination with the large, inter-generational households (*4*) could lead to infection fatality rates higher than those seen in high income countries (*1*). In addition, the high prevalence of comorbidities such as HIV/AIDS is predicted to lead to increased risk of severe COVID-19 in infected individuals (*5*). Moreover, the co-existence of infectious diseases such as malaria (*6*), tuberculosis (*7*), dengue (*8*), Ebola (*9*), and others (*10*) pose additional significant medical and infrastructure challenges in controlling the COVID-19 epidemic in Africa.

Meteorological variables have been linked to the transmission of and survival from seasonal influenza (*11–14*), severe acute respiratory syndrome coronavirus (SARS-CoV) (*15–17*) and Middle East respiratory syndrome coronavirus (MERS-CoV) (*18, 19*). It is therefore unsurprising that there are many recent studies exploring the link between temperature, humidity, and COVID-19. All studies to-date have focused on modeling, or on identifying a statistical link between meteorological variables against reported COVID-19 cases, without laboratory studies. A recent systematic review reports agreement among published research, with cold and dry conditions contributing to COVID-19 transmission (*20*). However, these results must be considered preliminary. Early studies of COVID-19 transmission focused on the emerging pandemic during the boreal spring (March through May), when the majority of cases were found in China, the US and Europe. It is difficult therefore to extrapolate meteorological results to the very different climates found in the tropics, for example from the recent outbreaks in India and Brazil. Many low and middle-income countries (LMICs), as defined by the World Bank, are located in the tropics, where many potential confounding factors which could mimic a weather signal. These factors include median age, testing and health capabilities, population density, access to sanitation and the number of new cases arriving in a country through global travel hubs (*21*). It is also difficult to extract seasonality from a single outbreak that began in the boreal spring, and considerable caution has been raised regarding tropical case-load and confounding factors (*22*).

The human response to the pandemic can also drastically shape its timing and intensity. Where data on social distancing is sparse, government testing and stringency policies (See Methods Section 4.3) can be used as a common surrogate to compare countries’ efforts to contain the spread of the virus, bolster healthcare systems, enact rigorous testing policy, and provide economic support. The Oxford Coronavirus Government Tracker (OxCGRT) standardizes these complex systems into a set of policy metrics in each of these domains (*23*). More strict social policies identified in the OxCGRT have been associated with reductions in human mobility (*24, 25*). Across 161 countries, some of these policies were significantly associated with lower per capita mortality (*26*) including: school closing, canceling public events, and restrictions on gatherings and international travel. Likewise, others have found that strict policies are negatively associated with the growth of new cases (*27–29*). The relationship between policy and observed changes in social distancing, case numbers, and mortality is complicated by an unknown delay of effect. One estimate indicates a decline in growth of new cases within one week of enacting strict policy, and deceleration of growth within 2 weeks (*29*). Although the implementation of containment policies can be and have been used by many African nations (*30*), lockdown cannot be maintained in these countries without a worsening of severe poverty and resultant loss of life (*31, 32*).

We have therefore developed a COVID-19 surveillance strategy that explores a growing spatiotemporal database on coronavirus epidemiology, meteorology, and social policy interventions. To model the spread of COVID-19 in Africa, we employ a data-driven endemic-epidemic model (*33*) to 1) visualize the burden of cases including the proportion of cases arising from sources local within-country and external between-country, 2) describe the factors which most correlate with spread, and 3) enable short-term forecasting of new cases. This modeling framework has been used previously to fit space-time dynamics of COVID-19 in Italy (*34*), Germany (*35*) and the United Kingdom (*36*) and to analyze other infectious diseases (*37*). The model is divided into three main parts: two epidemic components that capture sources of infections coming from within the country and from neighboring areas, and an endemic component that includes all contributions to the reported number of cases that are not taken into account by the epidemic part. The epidemic part of the model has an auto-regressive nature, this means that the past number of COVID-19 cases reported both within a specific country and in the rest of the continent will be used to forecast the present and future trend of COVID-19 cases. How much the past observations contribute to the the future disease count depends on two parameters, *λ* for the local transmission and *ϕ* for the external transmission and will be estimated from the data. In particular, the impact of cases reported in the neighboring countries depends also on a set of weights that modulate the spatial connectivity of the countries in the continent (see Method section). These two parameters are also functions of social policy, testing availability, meteorological and demographic factors whose association with transmission we aim to determine.

## 2 Results

### 2.1 COVID-19 spread and response

As of August 13, 2020, the 55 AU Member States had reported over 1,000,000 cases and 20,000 deaths from COVID-19. The southern region had the most cases, reporting over 50 percent (over 560,000 cases and 11,000 deaths) of the total for the continent. North Africa carries the highest regional case-fatality rate (4 per cent) but contributes 20 per cent of the continent’s cases, with countries such as Egypt (102 cases per 100,000), Morocco (94 cases per 100,000) and Algeria (83 cases per 100,000) driving the overall numbers (Figure 1). As more countries conduct targeted mass screening and testing, these figures continue to change. The spatial distribution of cases per 100,000 displays no clear geographical pattern (Figure 1). South Africa, Djibouti, Equatorial Guinea, Gabon and Egypt carry the largest burden of cases per capita, ranging from 100 to 500 per 100,000. The epidemiological curves for the African countries display varying shapes, mostly driven by the frequency and intensity of testing. For example, the epidemiological curve of South Africa is similar to those of the UK and the US (Supplementary Figure S1). An exception is Tanzania, which stopped reporting new cases in late April (Supplementary Figure S1).

**Figure 1:**
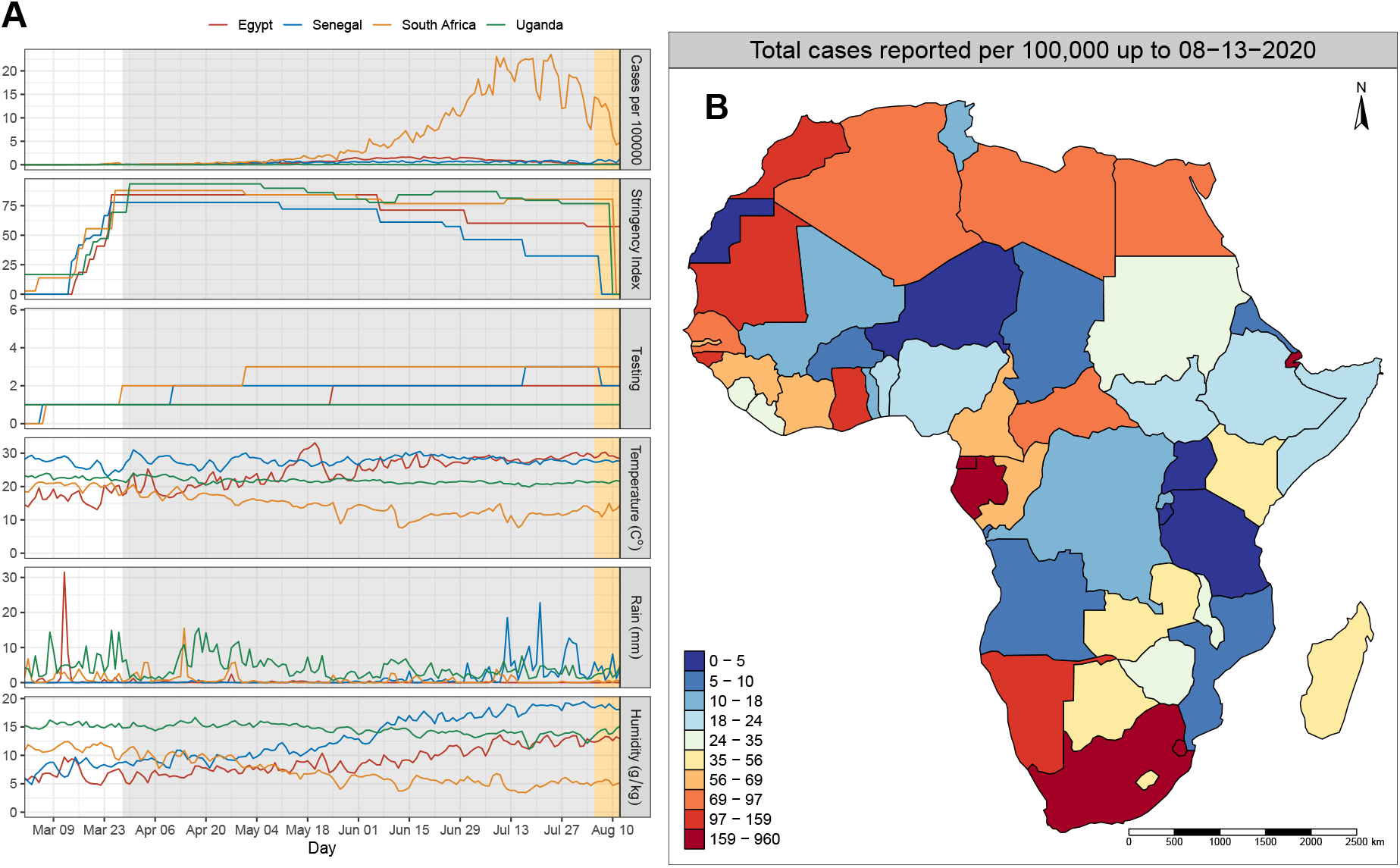
Temporal distribution of reported cases, stringency index, testing policy and weather factors. (A) Time series (February-August 2020) of (from top to bottom) daily reported cases per 100,000, stringency index, testing policy, temperature, rainfall, and specific humidity for representative countries Egypt, Senegal, South Africa and Uganda. These time-dependent covariates were used as predictors (explanatory variables) in the best-fitting model shown in Table 1. The gray and orange shaded areas show the time window of data used to fit the model and the data held-out for model validation, respectively. (B) Country-specific distribution of cumulative cases per 100,000 on August 13, 2020

Time series for case incidence and temporally-varying model inputs are shown for selected countries in the left panel of Figure 1. The full set of case incidence time series for all countries can be found in Supplementary Figure S1. A majority of the countries imposed containment policies, including lockdowns and curfews, to prevent further COVID-19 transmission within their borders in early March. These social policy interventions have remained in effect through August for most countries (Supplementary Figure S2). Testing policies, which were restrictive at the beginning of the pandemic due to inadequate testing infrastructure, have become more open as testing is made widely available (Supplementary Figure S3). As expected, spatiotemporal distribution of temperatures, rainfall and specific humidity are very heterogeneous across the continent (Figure 1, and Supplementary Figures S4, S5, & S6). Population weighted averages of these three meteorological variables were calculated for each country and day. This type of weighting prioritizes the human-climate interaction over the land-climate interaction (Supplementary Figures S7, S8, & S9).

**Table 1:** Maximum likelihood estimates and corresponding 95% confidence intervals for a model with 7-day lag. For climatic variables, a 1 standard deviation increase in climatic variables results in the shown relative risk. For stringency index, a 10% increase in stringency is associated with the increased relative risk shown. HDI and testing policy are on ordinal scale 0 to 3 (HDI) and 0 to 4 (testing policy). **Bolded** estimates are statistically significant. The spatial weight decay, *ρ*, reflects the strength of inter-country connectivity, and overdispersion parameter, *ψ*.

| Parameter | Final Model |  |  |
| --- | --- | --- | --- |
|  | Relative Risk | 95% CI | p-value |
| **Endemic** |  |  |  |
| Intercept | 11.071 | (7.150, 17.142) | - |
| **Within-country** |  |  |  |
| Intercept | 0.958 | (0.550, 1.669) | - |
| log(population) | 1.036 | (0.956, 1.123) | 0.391 |
| HDI | 0.957 | (0.819, 1.118) | 0.580 |
| Landlocked | **0.674** | (0.531, 0.857) | 0.001 |
| Stringency<sub>$t-7$</sub> | **1.872** | (1.170, 3.000) | 0.008 |
| Testing<sub>$t-7$</sub> | **0.817** | (0.729, 0.918) | 0.001 |
| Rain<sub>$t-7$</sub> | 1.045 | (0.981, 1.112) | 0.175 |
| Temperature<sub>$t-7$</sub> | **1.106** | (1.014, 1.206) | 0.023 |
| Humidity<sub>$t-7$</sub> | **0.856** | (0.780, 0.940) | 0.001 |
| **Between-country** |  |  |  |
| Intercept | 0.045 | (0.004, 0.481) | - |
| log(population) | 1.428 | (0.923, 2.210) | 0.110 |
| HDI | 1.239 | (0.584, 2.628) | 0.576 |
| Landlocked | 0.844 | (0.306, 2.323) | 0.742 |
| Stringency<sub>$t-7$</sub> | 1.630 | (0.560, 4.749) | 0.380 |
| Testing<sub>$t-7$</sub> | **2.322** | (1.676, 3.218) | < 0.0001 |
| $\rho$ | 2.186 | (1.532, 2.839) | - |
| $\psi$ | 1.703 | (1.631, 1.775) | - |

### 2.2 Optimal model

The model specification reported in Equations 3, 4 and 5 representing the endemic, within- and between-country component of the model, respectively, is the result of a model selection procedure based on the Akaike Information Criterion (AIC) (*38*). A summary of model comparison and selection process is presented in supplementary Table S1. We began with an intercept only model (Model 1) with a population offset in the the endemic component of the model and country’s measure of connectivity based on a power law. More complicated versions of the epidemic component were evaluated by sequentially adding weather, demographic, stringency index and testing policy in the formula for the within (*λ*) and between-country (*ϕ*) components of the model. We also tested whether multiple lags for cases and covariates better described the observed patterns: considering lags d = 1, …, D where D = 14 days. Exploration of the higher order model with Poisson and geometric lag weights, revealed that relative to the first-order model, the largest improvement in AIC (ΔAIC = -1560) was achieved with a model with D = 7 (Supplementary Figure S10). Therefore, the model with lag 7 days for cases, testing policy, stringency index including human development index (HDI), landlocked status and population in the between-country component, and meteorology factors, HDI, landlocked status, stringency index and testing policy in the within-country component, yielded the lowest AIC (from 48824.32 in model 1 to 48437.99 in model 5 without random effects, Supplementary Table S1).

Due to the high spatiotemporal heterogeneity of reported cases across Africa and to better capture country specific transmission dynamics and incidence levels not explained by observed covariates, we allowed the intercept (mean levels of *λ* and *ϕ*) in the local (4) and neighbor-driven (5) sources of infections to vary for each country. The relative risks for each explanatory variable included in this final model and the associated 95% confidence intervals are reported in Table 1. Landlocked status, stringency index, and testing policy were significant contributing factors on the local transmission of cases (Figure 2).

**Figure 2:**
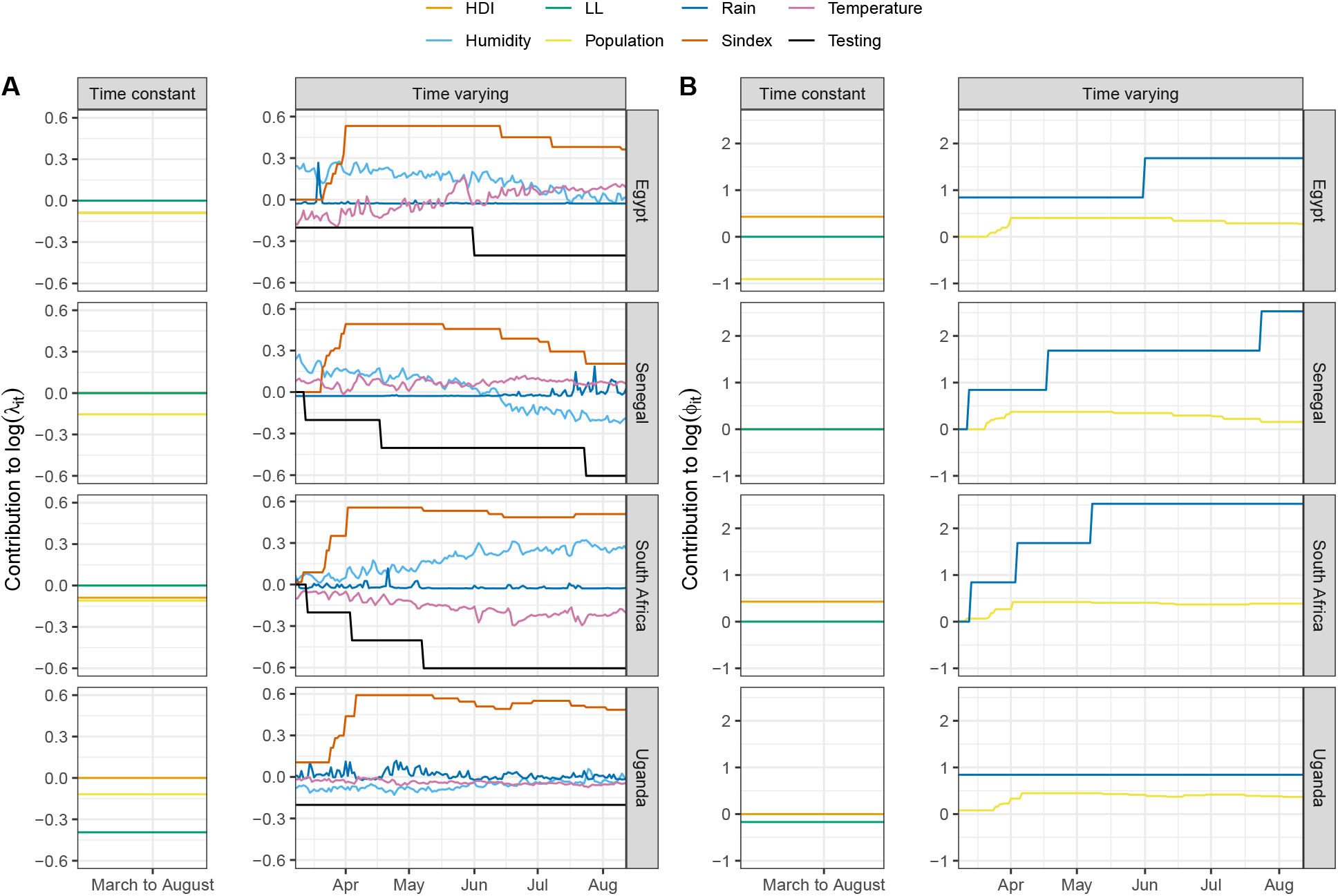
Magnitude and direction of the explanatory factors to the within and between country transmission. Time constant and time varying covariates from 4 selected countries showing variations in the strength and the direction of the contributions to the local within-country (A) and external between-country transmission of cases (B).

In addition, higher lagged mean temperature was a positive contributing factor but higher specific humidity had a negative effect on the transmission of cases. For example, a 1 standard deviation increase in the lagged mean temperature results in 11% higher contribution on the within-country transmission [p = 0.023, RR 1.11, 95% CI: 1.01 to 1.21]. However, a 1 standard deviation increase in the 7-day lag mean specific humidity resulted in a 14% lower contribution on the local transmission of cases [p = 0.001, RR 0.86, 95% CI: 0.78 to 0.94]. More accessible testing remained the only significant contributing factor explaining the numbers of cases from the neighboring countries. With each level increase in the openness of the testing policy from 0 to 4, the contribution to the transmission of cases from neighboring countries was higher by 2-fold (p*<*0.0001). The overdispersion parameter decreased from the fixed effect model (1.95, 95% CI: 1.87 to 2.03) to random effects model (1.70 95% CI: 1.63 to 1.78) as a sign that the random effects absorbed part of the unexplained variability between countries.

Figure 3 shows countries specific (random) effects for the local and neighbor components of the model. A value higher (lower) than 1 means that a country has an average transmission rate that is higher (lower) than the rest of the continent. This may be interpreted as a country-specific propensity to generate more or fewer cases given the past number of reported infected individuals. South Africa and Djibouti are the only African countries with an effect significantly higher than 1. On the other hand, the Republic of Congo is the only country with a significantly lower than the continental-level mean caseload. With respect to the between-country contributions to transmission of cases, Benin, Cameroon, Central African Republic, Ethiopia, Gabon, Ghana, Guinea, Malawi, Republic of Congo and Senegal had a significantly higher than continental-level mean number of cases. Angola, Chad, Lesotho, Namibia and Tanzania had lower cases means compared to the continental-level mean. Panels A and B of Figure 3 also show some interesting spatial clustering of these effects. The estimated variation of these country-specific effects in the within-country component of the model is small 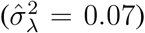 compared with their variation in the neighborhood component 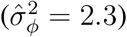. Although the between-area variability of transmission resulting from cases reported outside of the country was larger, the between-country intercept (Table 1) is very small and so the neighborhood component is in general a small contributor to the fit.

**Figure 3:**
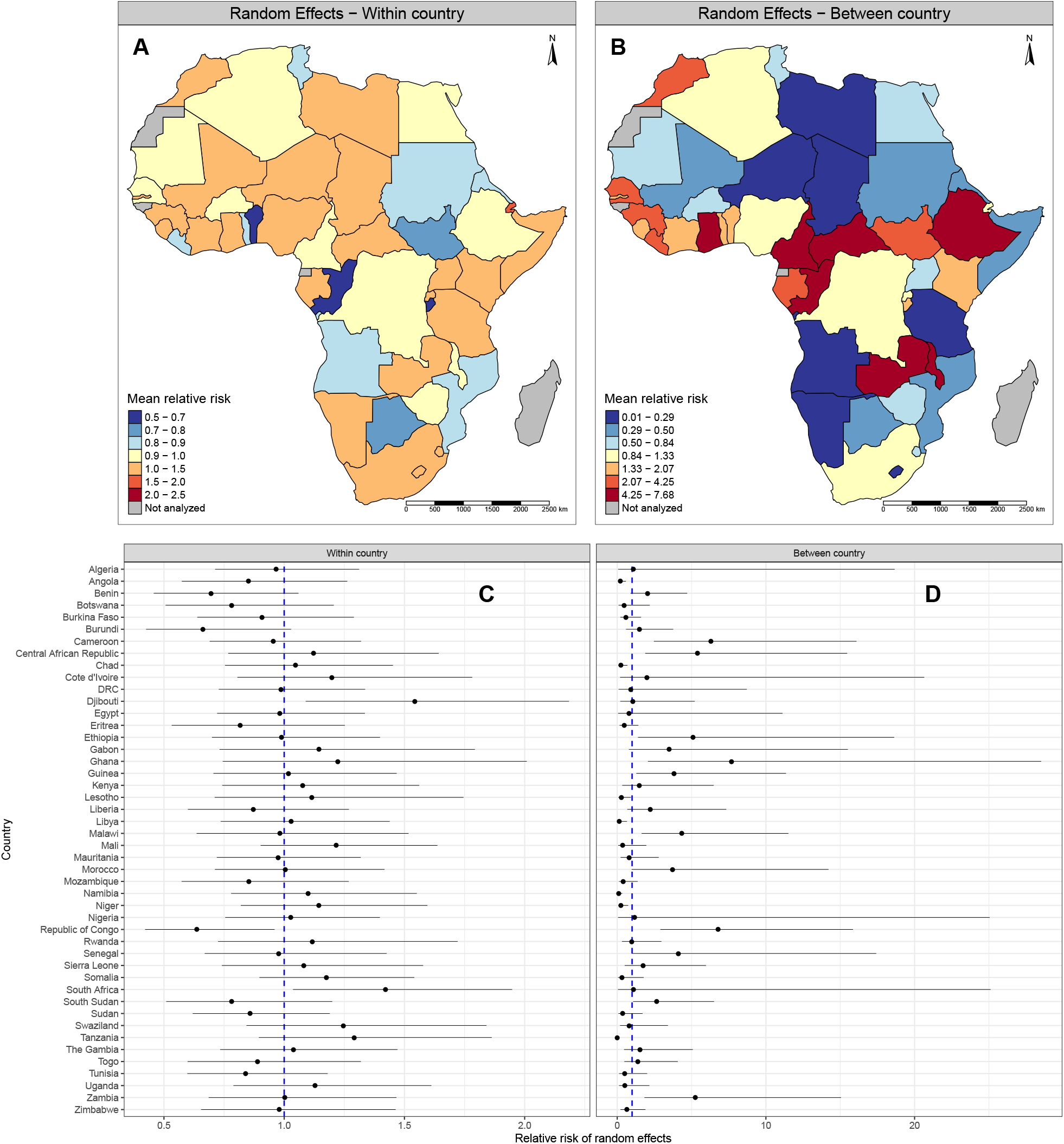
Random effects for the local and neighbor components of the model. Country-specific relative risks (RR) and their 95% CI for (A and C) within-country and (B and D) between-country model contributions. The dashed blue line in the forest plots represent continent-level average. RR greater than 1 indicates higher propensity for transmission as compared to the rest of the continent.

### 2.3 Contributions of within- and between-country transmission

We distinguish between endemic, within-country, and between-country contributions to the mean number of cases. Fitted values for all components according to model formulations in equation 1 are shown in Figure 4, with a complete listing in Supplementary Figure S11. The number of cases attributed to within- and between-country transmission of cases during the entire study period varied greatly. Across countries, the contribution from the endemic component was found to be minimal. Of the 46 countries analyzed, 16 of them are landlocked, and 13 (81%) of these had a substantial contribution of cases from their neighboring countries: Botswana, Burkina Faso, Burundi, Central African Republic, Ethiopia, Lesotho, Malawi, Rwanda, South Sudan, Swaziland, Uganda, Zambia, and Zimbabwe.

**Figure 4:**
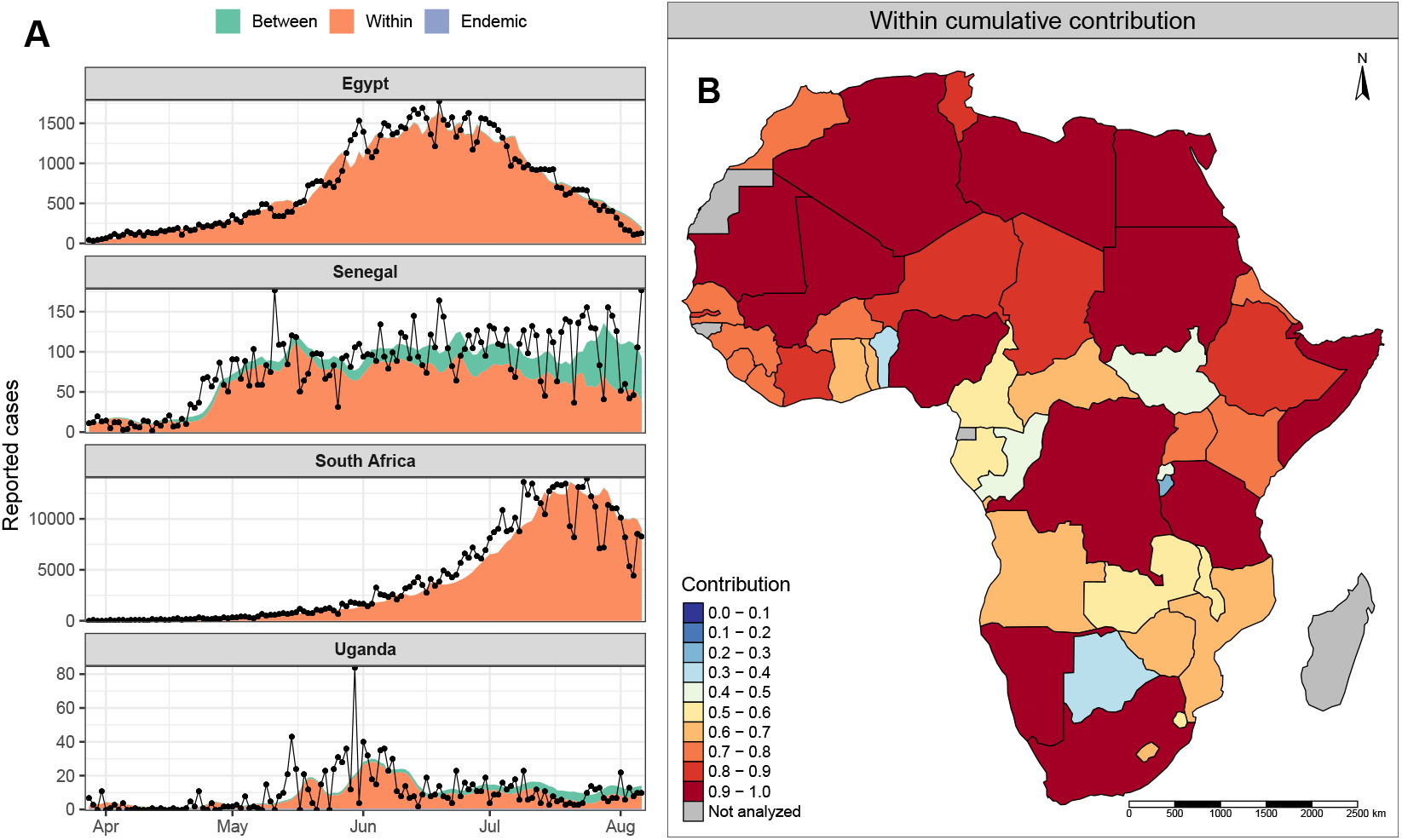
Contributions of new cases from endemic, within-country, and between-country model components. (A) Black dots and connecting lines are the observations and shaded colors are the model predictions from the three contributing components of the model. (B) The relative contribution of new cases from within-country sources, by country.

### 2.4 Short-term forecast

We keep the last 7 days of data out of the fitting procedure in order to use them as a forecast validation data set. We produce one-week ahead predictions and compare them with the reported data to check the quality of model forecast. The results show that the majority of individual country case count data are captured well within model prediction intervals (Figure 5 and Figure S12). Across countries, the model predictive performance was assessed with a calibration test based on proper scoring rules as described in (*39*). A map of p-values for the calibration test is shown in Figure S13. Overall model predictions are well calibrated and a misalignment between forecast and observations was only detected for few countries (*p <* 0.05, Burundi, Cameroon, Somalia, Botswana).

**Figure 5:**
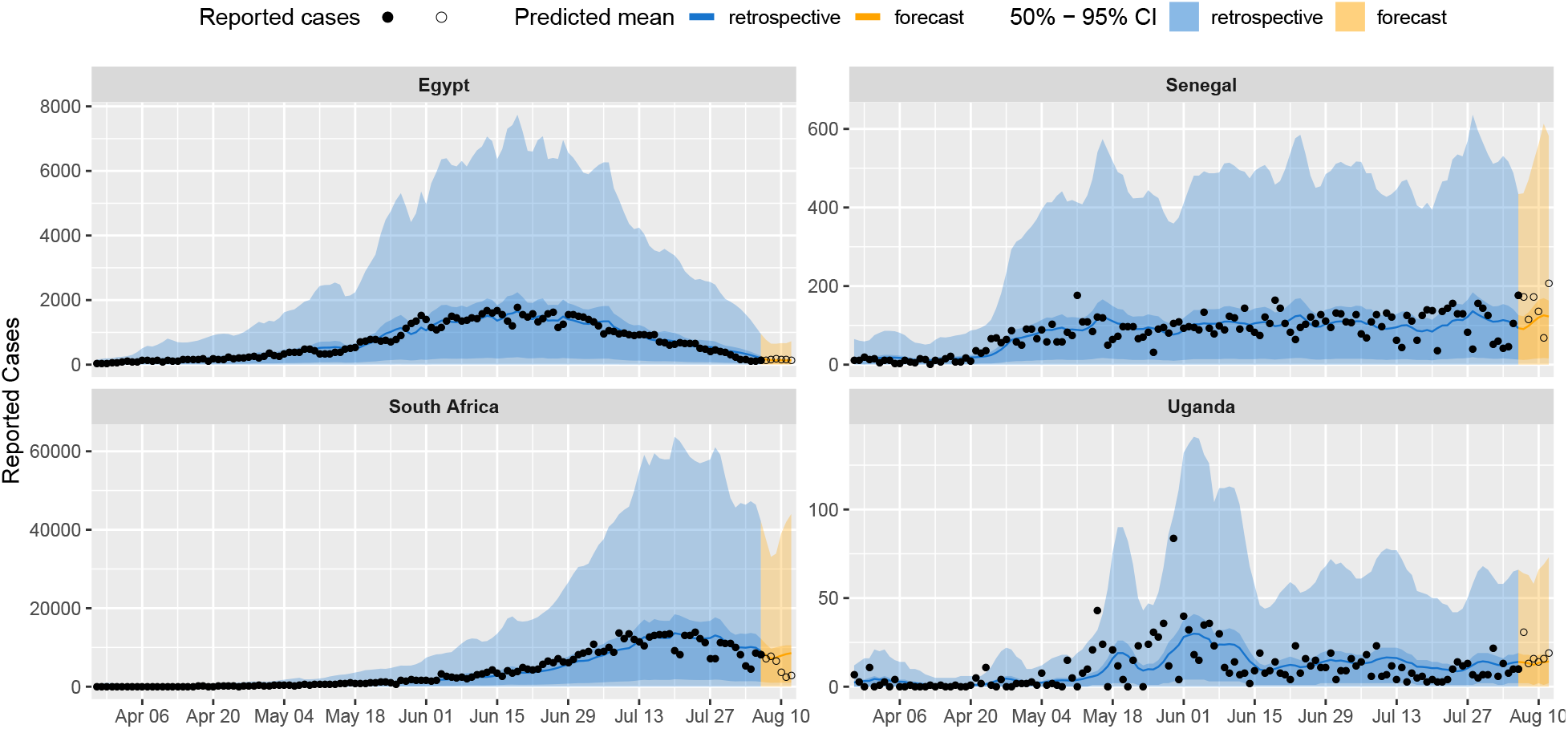
Retrospective and forecast model fit for selected countries. Retrospective model fit (blue shade) and forecast (orange shade). The 50% and 95% confidence intervals are represented by dark and light colors respectively. Filled circles in the blue shade are observed cases and the solid blue line the predicted retrospective mean. Open circles in the orange shaded area represent cases from model forecast and the solid orange line the predicted forecast case mean. Majority of individual country case count data are captured well within model prediction intervals. For the full list of countries see Figure S12.

## 3 Discussion

We present a model that can improve the ability of African countries to interpret the complex data available to them during the COVID-19 pandemic. This approach balances the simplicity and consequent robustness of an empirical model against the more complex, potentially more realistic but also more strongly assumption-driven kind of compartmental mechanistic models (*40*). A key feature of our approach is the ability to distinguish between case incidence arising from the local within- or neighbor-driven transmission of infection. Distinguishing within- and between-country transmission of cases allows us to identify potential strategies for social or health policy intervention. The model further enables reproducing the history of the epidemic in relationship to past policy, and producing short-term predictions of the dynamic evolution of the epidemic.

We find that a country’s testing capacity, social policy, landlocked status, temperature and humidity are important contributing factors explaining the within and between-country transmission of cases. The availability of more testing to a wider swath of the populace is a potent contributor for reduced case transmission within country, while having the opposite effect on case transmission from neighboring countries. Testing policy, another surrogate for healthcare capability and preparedness to handle the pandemic, demonstrates this unique opposing effect on the two model components. Countries in northern and southern Africa that have relatively high HDI demonstrated comparatively higher numbers of cases per population. On the other hand, even in the face of border closures, landlocked countries depend on open borders for trade. For such countries, strict border closure measures are difficult to impose, enabling a constant influx of cases from the neighboring countries.

The observed association of temperature and specific humidity with the case numbers, although small, points to the possible biological and behavioral responses to weather patterns, which in turn drive the dynamics of SARS-CoV-2 infection. Temperature and humidity are known factors in SARS-CoV, MERS-CoV, and influenza virus survival (*41–43*). Lower humidity has been consistently associated with higher cases. Besides potentially prolonging half-life and viability of the virus, other potential mechanisms associated with low humidity include stabilization of the aerosol droplet, enhanced propagation in nasal mucosa, and impaired localized innate immunity (*44*). Whether the observed association is driven by the change in social behavioral patterns or the effect on the survival of SARS-CoV-2 remains to be explored (*45*). It is also possible that the observed contribution of meteorological factors to case transmission might be an artifact of spatial averaging and assigning one meteorological value to an entire country. It will be important to explore such associations in more detail before any policy relevant conclusions can be drawn. Thus, at present, policy makers must focus on social-behavioral interventions such as reducing physical contact within communities, while COVID-19 risk predictions based on climate information alone should be interpreted with caution (*46*).

Our infection surveillance tool adds to the public health capacity already in place on the continent to better understand transmission patterns between and within African countries. Containment and mitigation strategies to limit the spread of the virus, including restrictions on movement, public gatherings, and schools, were implemented very early in the pandemic. In a resource-limited setting such as Africa, containment and mitigation strategies remain the most robust defense against high infection rates and mortality. However, it is anticipated that physical-distancing measures enforced to limit transmission will also restrict access to essential non-COVID-19 healthcare services, such as disruptions in the existing programs for tuberculosis, HIV/AIDS, malaria, and vaccine-preventable diseases, causing long-lasting collateral damage on the continent (*32*). Although, between 29 million to 44 million individuals (*2*) in Africa are projected to become infected in the first year of the pandemic if containment measures fail, these numbers may be underestimated since the proportion of asymptomatic infections is not well established. Since detection is biased towards clinically severe disease, the attack rate of the infection is probably substantially higher than what is reported. At the beginning of the pandemic, it was estimated that up to 86% of all infections were undocumented and were the source of 79% of the documented cases (*47*). Such observations explain the rapid geographic spread of the infections and challenging efforts at containment. The number of asymptomatic cases is best determined by population-based seroepidemiology data. However, due to the fragile healthcare systems of Africa countries, this type of disease surveillance remains limited. On the other hand, it is also plausible that the lower incidence rate of the virus in Africa is because of the investment in preparedness and response efforts toward various outbreaks on the continent (such as Ebola virus disease, Lassa fever, polio, measles, tuberculosis, and human immunodeficiency virus) (*32*). This technical know-how has been swiftly adapted to COVID-19.

An additional strength of our modeling strategy is the ability to incorporate the disease-specific serial interval between sequential infections in the autoregressive model. We attempted to mimic the longer (greater than 1 day) serial interval (*48, 49*), infectiousness (*48, 50*), and latency (*48*) of COVID-19 transmission, by extending the observational interval of the infectious process to several days. The Poisson autoregressive weighting method used in our modeling strategy also captures an initial increase in infectiousness and may thus be more appropriate for longer serial intervals or daily data. In their recent work, Bracher and Held show that moving beyond one day lags to higher order time lags improves predictive performance of these endemic-epidemic models (*51*). For our optimization scheme we tested lags up to 14 days, and found that a lag at 7 days provided the best model fit. Short-term predictions enable the monitoring of case incidence trends but are limited by high levels of uncertainty. This is the result of the non-negligible overdispersion detected in the data and due to the several sources of unmodeled spatial and temporal heterogeneity across the continent.

### 3.1 Limitations

A number of assumptions were made in our analysis. Contact patterns across countries were assumed to be constant over time. Current patterns, such as inter-country air travel and border crossing by land might not follow the weights we have assumed, and actual population contact probabilities might not be constant over time. Nevertheless, the use of higher order neighborhood (beyond sharing a border) contact patterns led to improved model fit compared to an assumption of first order neighbors (bordering countries) only. Other assumptions related to the testing and stringency policies. These are coarse approximations of governmental response to the surveillance and control disease transmission. The absence of quantifiable tests per capita is a limitation of this approach.

We are aware that SARS-COV-2 is often carried by apparently healthy individuals who might unknowingly transmit the pathogen. In the present analysis, we could not disentangle asymptomatic and symptomatic disease. Under-reporting can introduce artifacts in the autocorrelation structure and may confound the estimation of lag weights of the underlying serial interval distribution (*52*). Additionally, we assumed that model coefficients were constant over time. This is not the optimal fit for SARS-COV-2 transmission if there is seasonal variation, and implies that the interaction with weather is the same in summer and winter. We avoided adding sinusoidal (smooth, repetitive oscillation) functions in the endemic component because of the short interval of the current pandemic not spanning a whole year. Lastly, we excluded Equatorial Guinea, Guinea-Bissau and Western Sahara due to missing stringency index and the six island nations (Madagascar, Comoros, Mauritius, Seychelles, Cape Verde, São Tomé and Príncipe) due to the lack of connectivity to mainland Africa which prevented model convergence. This modeling strategy is limited by the quality of the data and the lack of non-linear dynamics in the model.

### 3.2 Conclusions

We present a pan-African COVID-19 surveillance tool to track and perform short-term forecast COVID-19 cases and to quantify between- and within-country sources. Our analyses give insight into the sociodemographic, geodemographic, testing, mitigation/containment and meteorological factors that influence the spread of the SARS-CoV-2 infection. Although our strategy can be used for short-term predictions of cases, their accuracy heavily depends on the quality of testing and reporting data. In settings with fragile health systems, coupled with the vulnerability of lower HDI economies, the capacity to effectively track the pandemic is especially challenging. Such challenges point to the potential advantages in regional efforts to coordinate resources to test and report cases. Seeking equitable behavioral and social interventions, balanced with coordinated country-specific strategies in infection suppression, should be a continental priority to control the COVID-19 pandemic in Africa.

## 4 Materials and Methods

### 4.1 Overview

Our analyses included 46 countries of mainland Africa. We do not provide estimates for Equatorial Guinea, Guinea-Bissau and Western Sahara due to the missing data on stringency index, or the six island nations (Madagascar, Comoros, Mauritius, Seychelles, Cape Verde, São Tomé and Príncipe) due to the lack of spatial connectivity. Modeling the spread of COVID-19 over the African continent poses challenges, given the extensive cultural, political and environmental heterogeneity between countries. Indeed, this heterogeneity results in substantial variability of reported case counts across countries. It is this variability in case counts that motivates our choice of a relatively simple data-driven autoregressive modeling approach. Such a modeling approach focuses on the interaction of cases reported in time and space without hidden variables to be estimated.

### 4.2 Meteorology/Weather Factors

The seasonality of influenza transmission has been associated with cycles of temperature, rainfall, and specific humidity, although in different regions of the world transmission may peak during the “cold-dry” season (temperate climates) or during “humid-rainy” season (tropical climates) (*53*).

We estimated the influence of meteorological factors on the transmission dynamics of COVID-19 in Africa. Real-time, daily in-situ synoptic weather observations are sparse across much of Africa. Therefore daily, 10km spatial resolution mean temperature, rainfall and specific humidity data were obtained from UK Met Office numerical weather prediction model output (*54, 55*). These data are extracted from the early time steps of the model following data assimilation, to more closely approximate an observational dataset. This approach also has the advantage that future studies have access to the same coherent dataset at a global scale for applications outside of continental Africa. The weather product that generates these data closely approximates an observational dataset at locations that have dense observation coverage, whereas in observation-sparse areas the dataset relies more heavily upon the numerical weather prediction model (a physics-based, rather than statistical model). The dataset contained no missing data.

A population density-weighted spatial average (supplementary Figures: S7, S8, S9) was then applied for each day and country using the R package **exactextractr** (*56*). Population density was obtained from the Gridded Population of the World version 4 (GPWv4) from the Centre for International Earth Science Information Network (CIESIN) (*57*). Weighting climate variables by population gives a closer approximation to the weather conditions faced by humans living in that country compared to an unweighted average over total land area. For example, the country of Algeria, in which much of the population resides along the coast, demonstrates a cooler, wetter, and more humid climate when weighting by population (Supplementary Figures S7E, S8E, & S9E).

### 4.3 Stringency index and testing policy

To include an aggregate measure of countries’ social policies, the *stringency index* sourced from the OxCGRT data set was used. This composite measure reflects government policies related to school and workplace closures, restrictions on public gatherings, events, public transportation, limitations of local and global travel, stay-at-home orders, and public education campaigns (*23*). A full description of these variables is provided in Supplementary Table S2. The stringency index is calculated from these categorical variables using a weighted average, with a range of 0 to 100 indicating weak to strict stringency measures, respectively. A time-dependent metric of testing policy was also extracted from this dataset. Ranging from 0 to 4 this categorical metric increases with more open and comprehensive testing policy.

### 4.4 Human Development Index, Demography, United Nations Geographic Regions and Coastline Access

In our modeling strategy we incorporate key socioeconomic and sociodemographic epidemiological data including human development index (HDI), population, United Nations geographic regions and coastline access (supplementary Figure S14). HDI represents the national data on key aspects of development, namely education, economy and health (*58*). The HDI is the geometric mean of normalized indices for each of the three dimensions. The education dimension is measured by average years of schooling for adults aged 25 years and more and expected years of schooling for children of school entering age. The economy dimension is measured by gross national income per capita, and the health dimension is assessed by life expectancy at birth. In Africa, majority of the countries fall in the low HDI category (supplemental Figure S14). The northern part of Africa and South Africa has a considerably higher HDI compared to the rest of the continent. Country-specific median age was correlated with HDI Pearson’s correlation coefficient (R=0.71, p*<*0.0001), Supplemental Figure S15, therefore we excluded this covariate from the model. We include in the model the 2020 population obtained from the Population Division of the Department of Economic and Social Affairs of the United Nations Secretariat (*59*). The categorization of sub-Saharan and Northern Africa was based on the United Nations geoscheme for Africa (*60*). This regional factor captures the human genetics (*61*), environment and climate (*62*), and sociocultural and sociodemographic variations of the African population (*63*). Finally, lack of direct access to the coastline may influence the flow of infections from neighboring countries as border trade remains an essential operation. For example, Uganda introduced border closures and tighter preventive measures on truck drivers’ movements during the epidemic; despite this, a substantial number of new infections have been imported from truck drivers crossing the border for trade (*64*). Such cross-border commerce remains a crucial part of the supply chain for landlocked African countries such as Uganda and Rwanda.

### 4.5 Model formulation

We chose a class of multivariate time series models for case count data introduced by Held et al (*65*), and further extended by Bracher and Held (*51*) with the addition of higher-order distributed lags.

New COVID-19 cases *Y*_*it*_ from country *i* at time *t* are assumed to be conditionally independent given past observations *Y*_*i,t*−*d*_, *i* = 1, …, *N, d* = 1, …, *D*, and distributed according to a negative binomial distribution with mean *µ*_*it*_ and overdispersion parameter *ψ* as

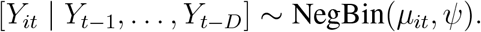

The conditional variance is 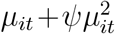, which demonstrates the role of the overdispersion parameter to capture variability greater than the mean. The conditional mean *µ*_*it*_ is decomposed into three additive components,

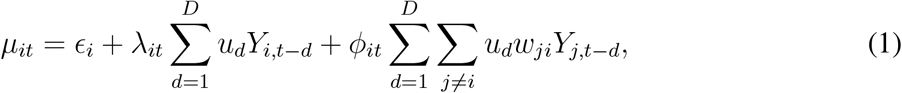

where *ϵ*_*i*_, *λ*_*it*_, and *ϕ*_*it*_ represent three contributions to case incidence. The first term, *ϵ*_*it*_, is the so-called *endemic* component and captures infections arising from sources other than past observed cases (e.g. contributions from areas that are not included in the neighbor set). The two other terms in (1), *λ*_*it*_ and *ϕ*_*it*_, constitute the *epidemic* part of the model and modulate how infective individuals reported in the past *d* days both locally and from neighboring countries will contribute to the average future number of reported cases. The strength of connection between countries is described by spatial weights *w*_*ji*_. This inter-country transmission susceptibility is defined using a power-law formulation proposed by Meyer and Held (*66*),

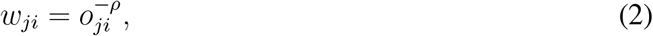

where *o*_*ji*_ is the path distance between countries *j* and *i* (with *o*_*ii*_ = 0, *o*_*ji*_ = 1 for direct neighbors *i* and *j* and so on) and *ρ* is a decay parameter to be estimated from the data. The spatial weights are normalized such that Σ_*k*_ *w*_*jk*_ = 1 for all rows *j* of the weight matrix (Supplementary Figure S16).

The normalized autoregressive weights *u*_*d*_ are shared between the local and global epidemic components, and represent the probability for a serial interval of up to *D* days - which is the average time in days between symptom onset in an infectious individual (or primary case) and symptoms appearing in a newly infected individual (or secondary case) when both are in close contact (*67*).

The parameters *ϵ*_*it*_, *λ*_*it*_, and *ϕ*_*it*_ are constrained to be non-negative and modeled as the natural log-transformed linear combination of different country-specific covariates. The endemic component,

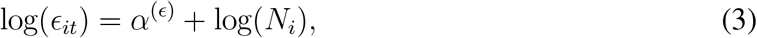

is decomposed as a constant *α*^(*ϵ*)^ specific to the baseline endemic and a term proportional to the country-level population *N*_*i*_. In the epidemic part of the model, we expect new cases to also be driven by country-specific factors: population (*N*_*i*_), Human Development Index classifications of low, medium, or high (*HDI*_*i*_ = {0, 1, 2}) and land-locked (*LL*_*i*_) status for each country are assumed constant over the time scale of analysis. Other forces driving new cases vary over time, either as a response to policy changes or natural fluctuation in environmental or societal patterns. Time-dependent covariates include mean daily temperature (*T*_*i,t*−*τ*_), rainfall (*R*_*i,t*−*τ*_), specific humidity (*H*_*i,t*−*τ*_), testing policy (*X*_*i,t*−*τ*_) and government stringency index (*S*_*i,t*−*τ*_), lagged at *τ* days. The full set of explanatory variables that contribute to the model from both internal and external epidemic components are formalized in (4) and (5) as

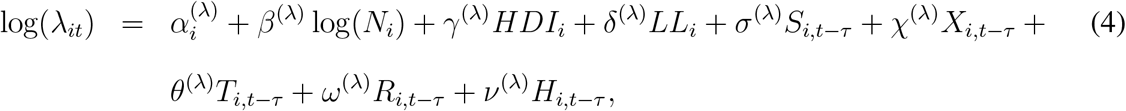

and

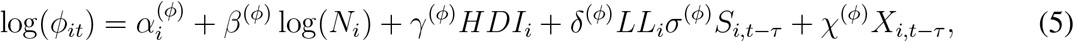

where 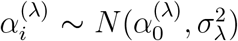 and 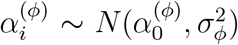 are a set of independent country-level random effects. This modeling framework is implemented in the R package **surveillance** (*68*). A complete table of data sources for model input is found in Supplementary Table S3.

#### Model fitting

We selected models based on Akaike’s Information Criteria (*69*) (AIC) if random-effects were not present (Supplementary Table S1). To compare models that included random effects we used proper scoring rules for count data (*70*). Scoring rules are functions *S*(*P, y*) that evaluate the accuracy of a predictive distribution *P* against an outcome *y* that was observed. We chose the model with the lowest AIC or with the lowest logarithmic score computed as minus the logarithm of the predictive distribution evaluated at the observed count. We began with the first-order autoregressive modeling (*D* = 1 in (1)) of daily COVID-19 incidence using intercept only model population offset and country connectivity. In a mechanistic interpretation of such a first-order model, the time between the appearance of symptoms in successive generations is assumed to be fixed to the observation interval at which the data are collected, here as one day (*52*).

After the estimation and illustration of this basic model, we expand the model by sequentially adding the following additional covariates: country-specific HDI, population in the both the within-country and neighboring countries, meteorology factors, stringency index, testing policy, landlocked status and random effects to more fully account for unobserved heterogeneity of the cases. Social policies and meteorological data were included in the model, testing for fit at different lags (for example, *T*_*i,t*−*τ*_, *τ* ∈ 0, 7, 14 days).

#### Model predictions

As in previous work by Held and Meyer, we use plug-in forecasts: forecast from the fitted model without carrying forward the uncertainty in the parameter estimates (*71*). We assess both the model fit and one-week-ahead forecast of the higher order autoregressive model with the logarithmic score. The smaller the score, the better the predictive quality (*72, 73*). Mean scores were generated for each country’s forecast, by averaging the log-score obtain for each day of the validation week.

## Data Availability

All data are in the public domain, and all code is open source and being made available at a public repository.

https://github.com/Schiff-Lab/COVID19-HHH4-Africa

## 5 Acknowledgments

We thank the Ugandan Ministry of Health, and the National Planning Authority for providing Uganda testing data.

## 6 Funding

This work was supported by a U.S. National Institutes of Health (NIH) Director’s Transformative Award 1R01AI145057 (SJS). MMN was supported in part by the Covid Virus Seed Fund from the Institute for Computational and Data Sciences and the Huck Institute at the Pennsylvania State University.

## 7 Author contributions

Conception and design: PS, CF, AG, MMN and SJS. Data acquisition: PS, CF, and AG. Statistical analysis and interpretation of the data: PS, CF, AG, HG, SJG, MMN, SJS. Drafting of the manuscript: PS, CF, AG, SJG, AJW, MMN, SJS. Critical revision for important intellectual content, discussion, review and final approval of the manuscript: all authors. Responsible for the overall content as the guarantors: PS, CF, AG and SJS.

## 8 Competing interests

All authors declare no competing interests.

## 9 Data and materials availability

All code and data are available in the supplementary materials and posted online at https://github.com/Schiff-Lab/COVID19-HHH4-Africa.

## 10 Supplementary Materials

- Code: Github code repository

**Figure S1:**
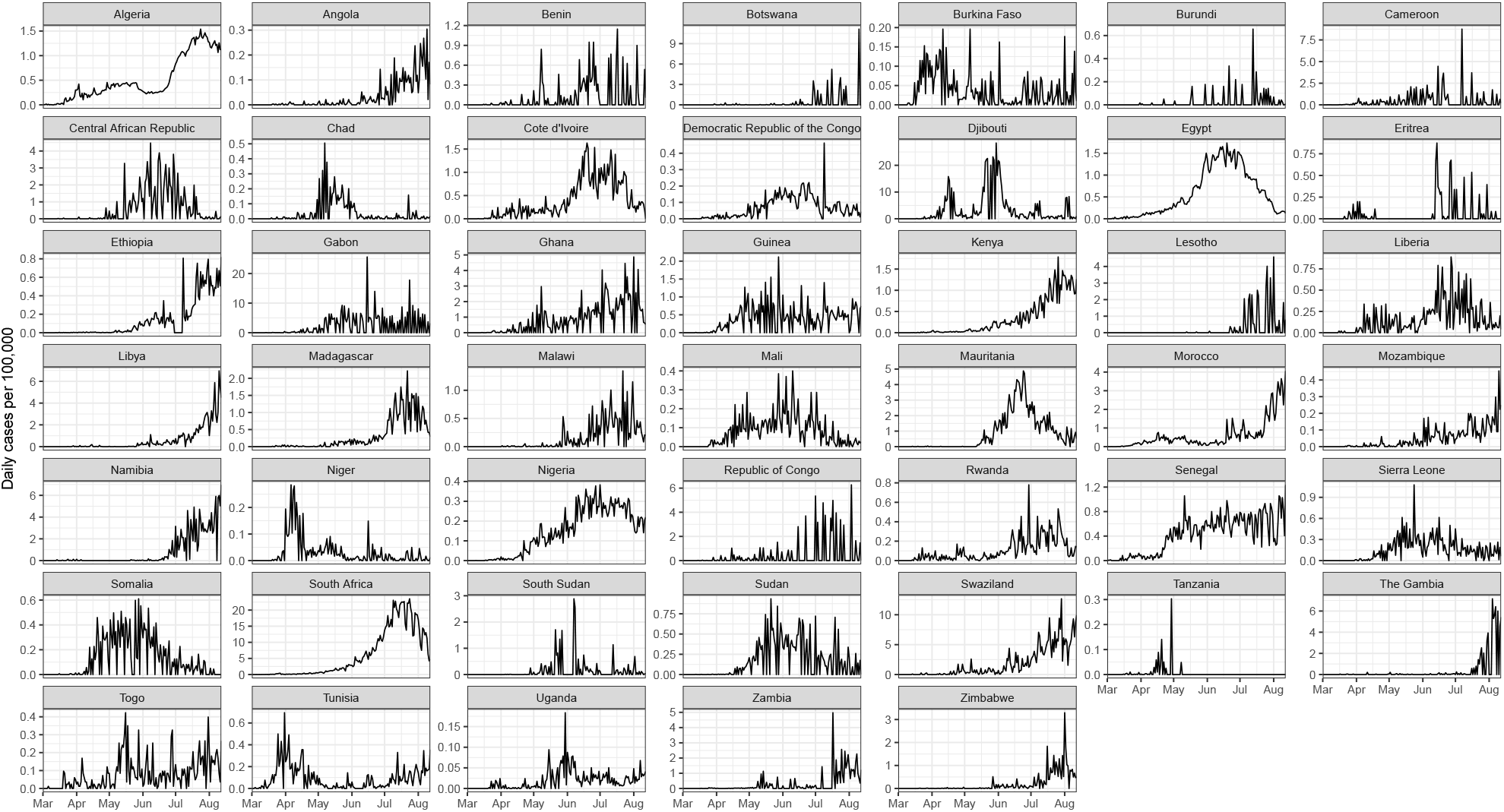
Time series of reported cases for all countries of Africa. Cases per 100,000 individuals between March 1 and August 13 are shown.

**Figure S2:**
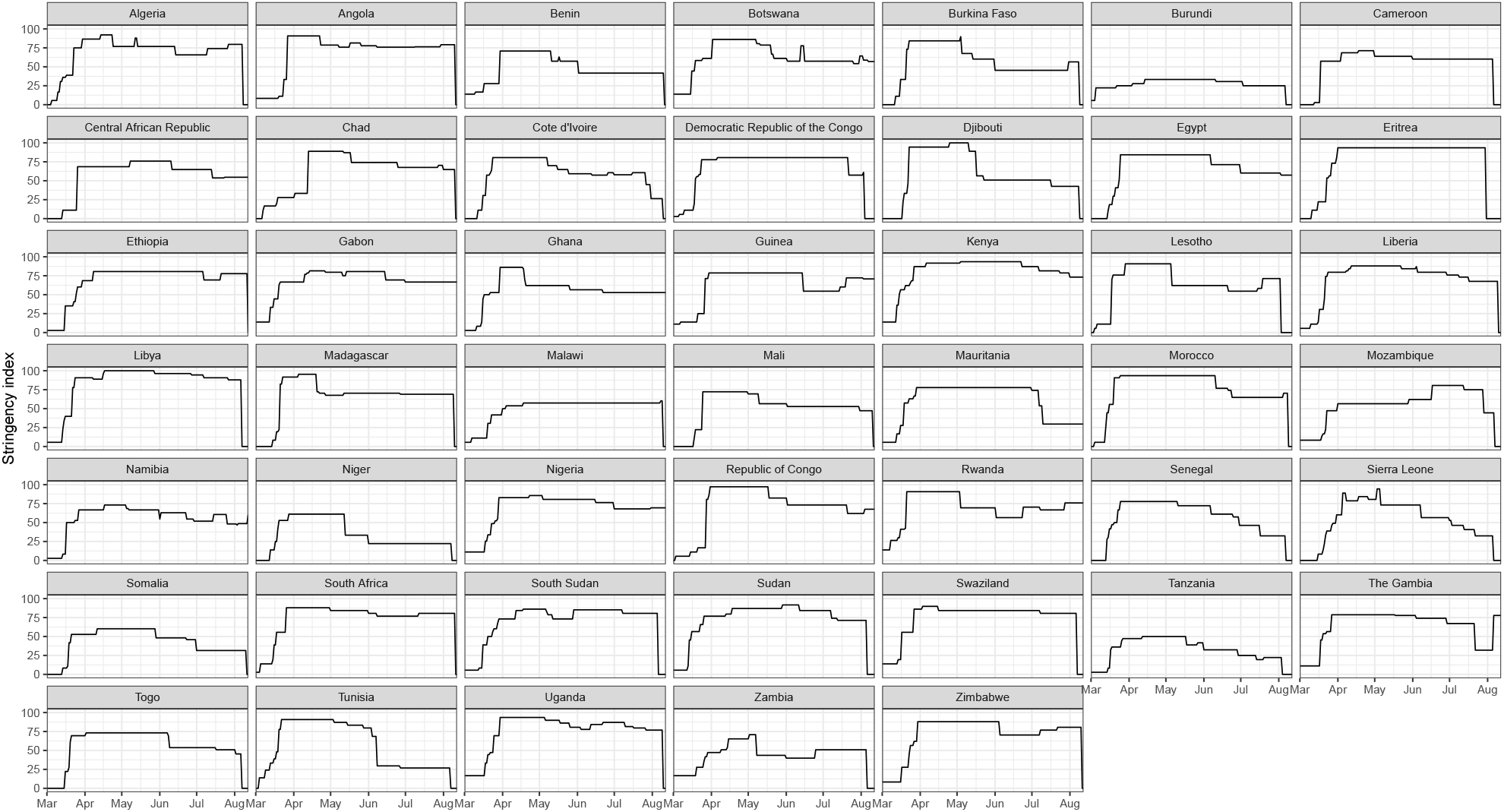
Time series of stringency index for all countries of Africa. This stringency index ranges from 0 (no government stringency policies) to 100 (very strict stringency policies).

**Figure S3:**
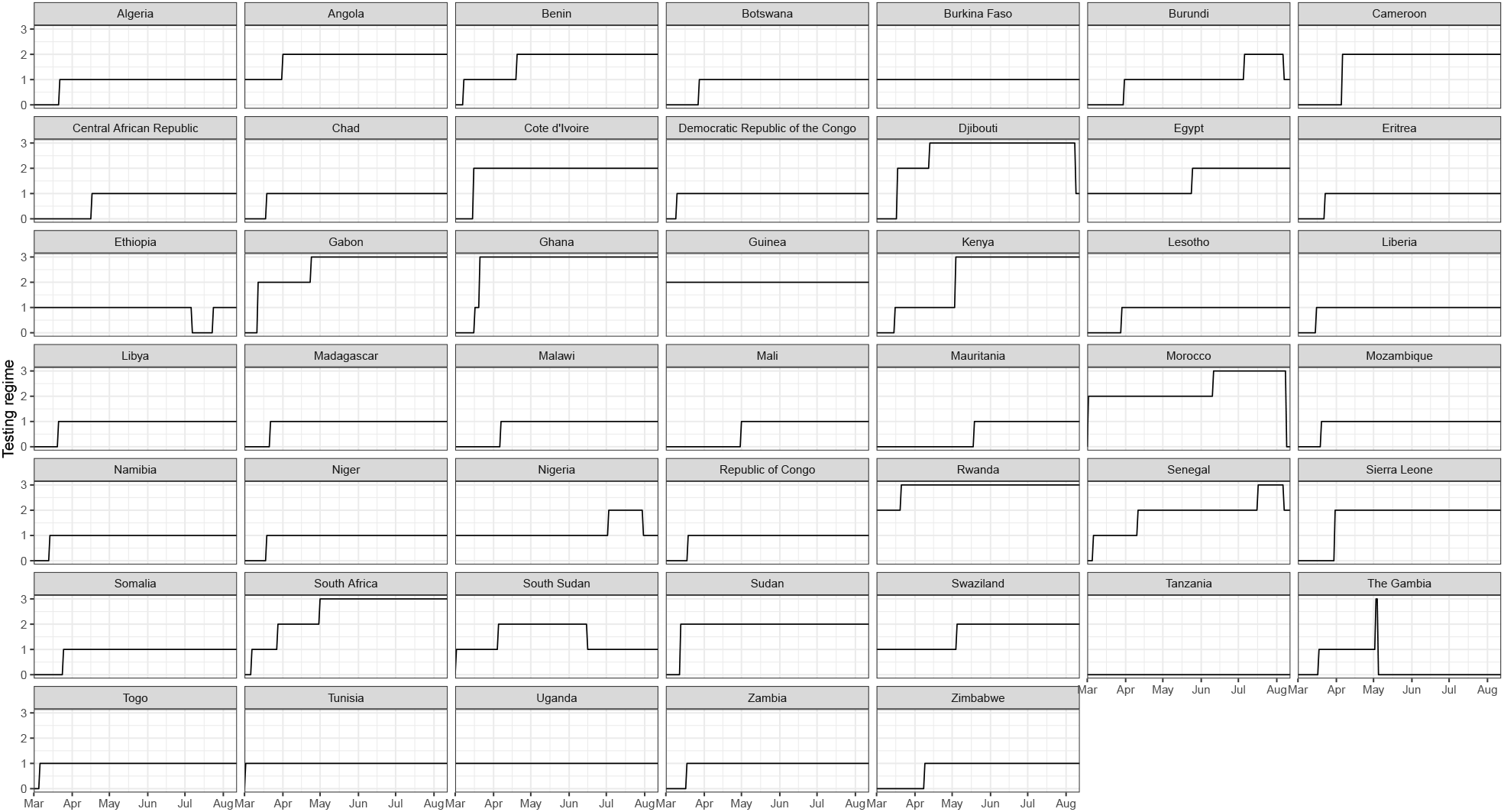
Time series of testing policy for all countries of Africa. This categorical variable (H2 in the OxCGRT codebook) ranges from 0 to 3, reflecting: 0-no testing policy, 1-testing those meeting certain criteria, 2-testing of anyone showing COVID-19 symptoms, and 3-open public testing.

**Figure S4:**
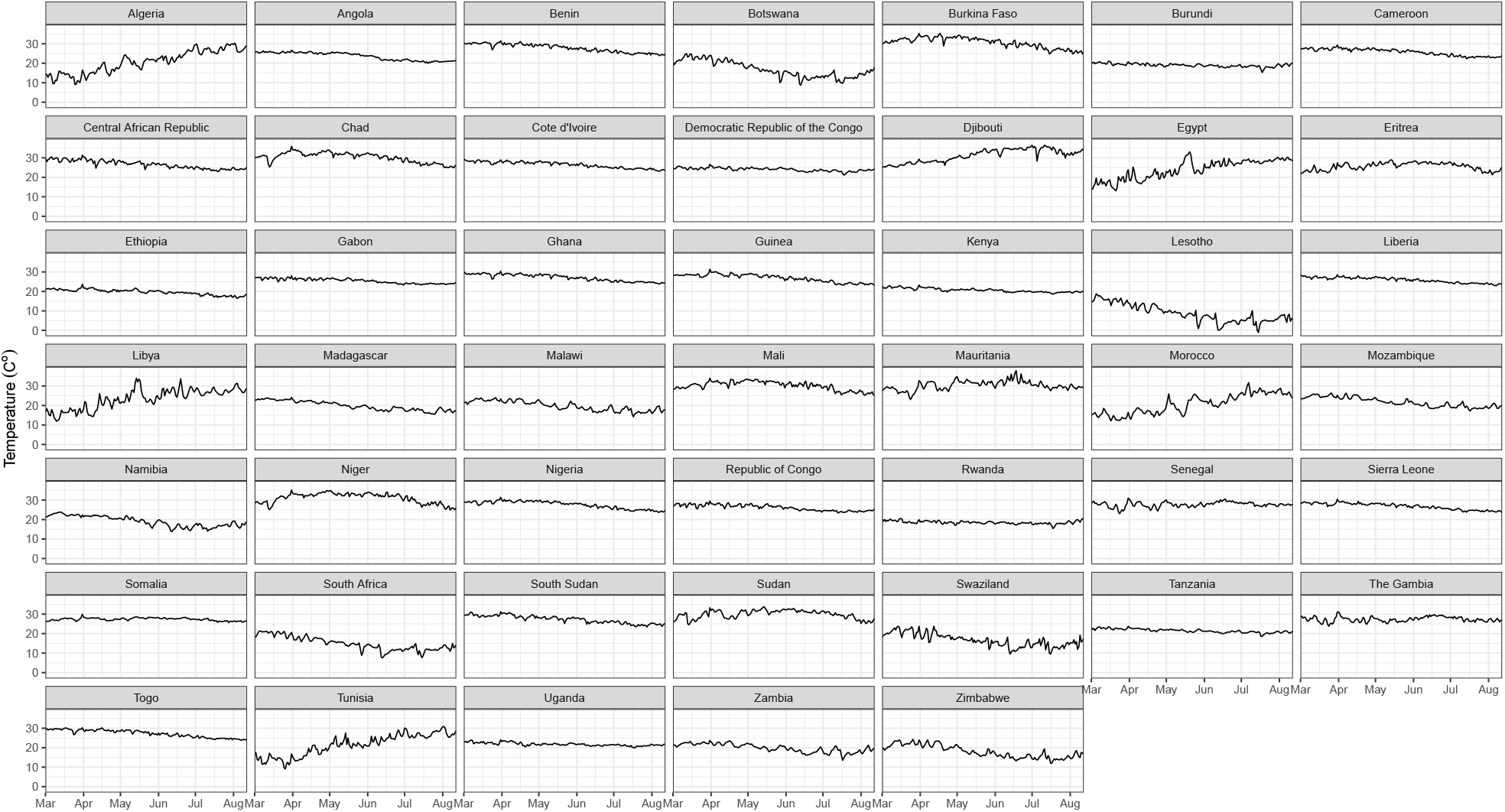
Time series of population-weighted mean temperature for all countries of Africa.

**Figure S5:**
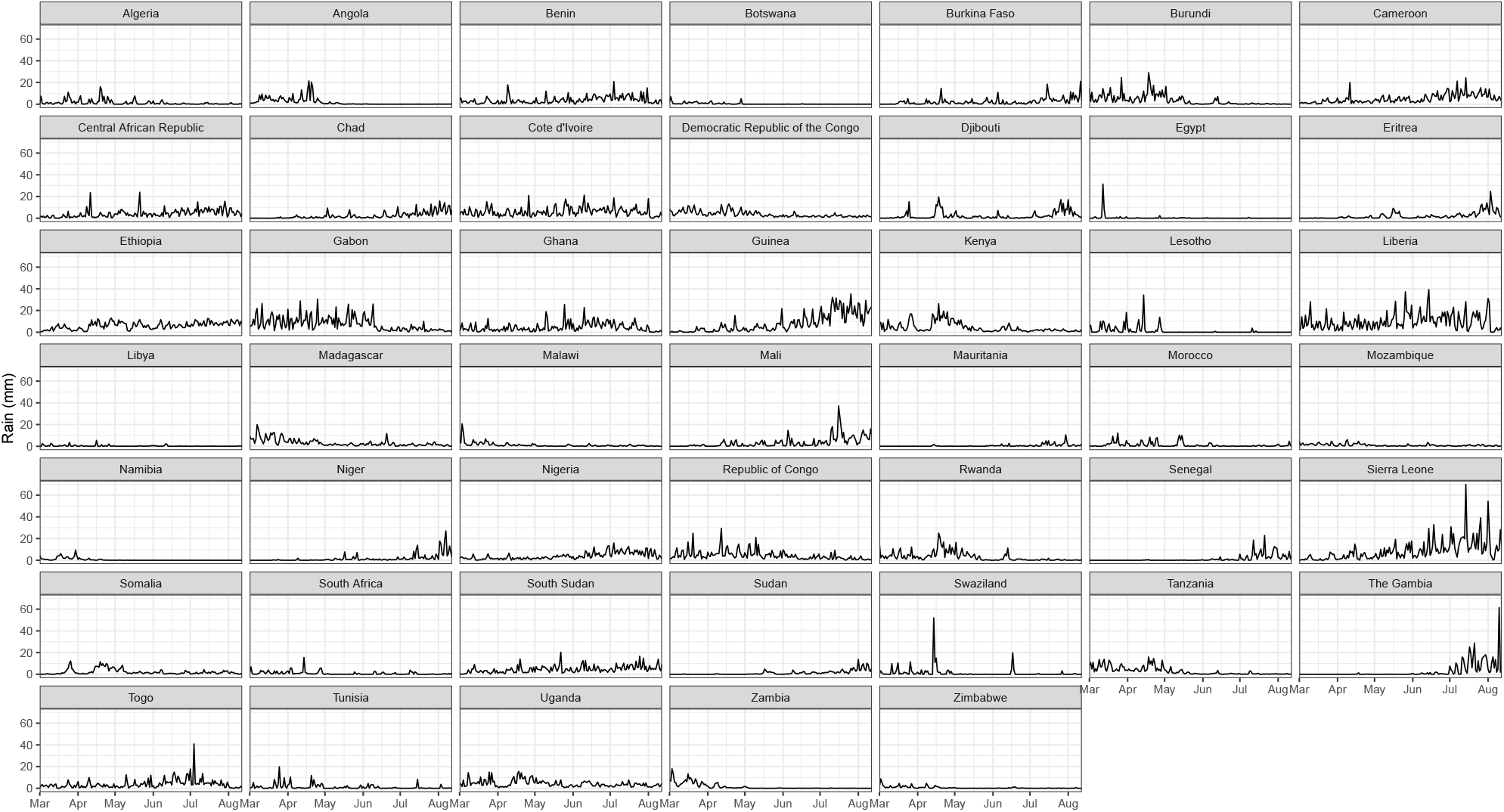
Time series of population-weighted mean rain for all countries of Africa.

**Figure S6:**
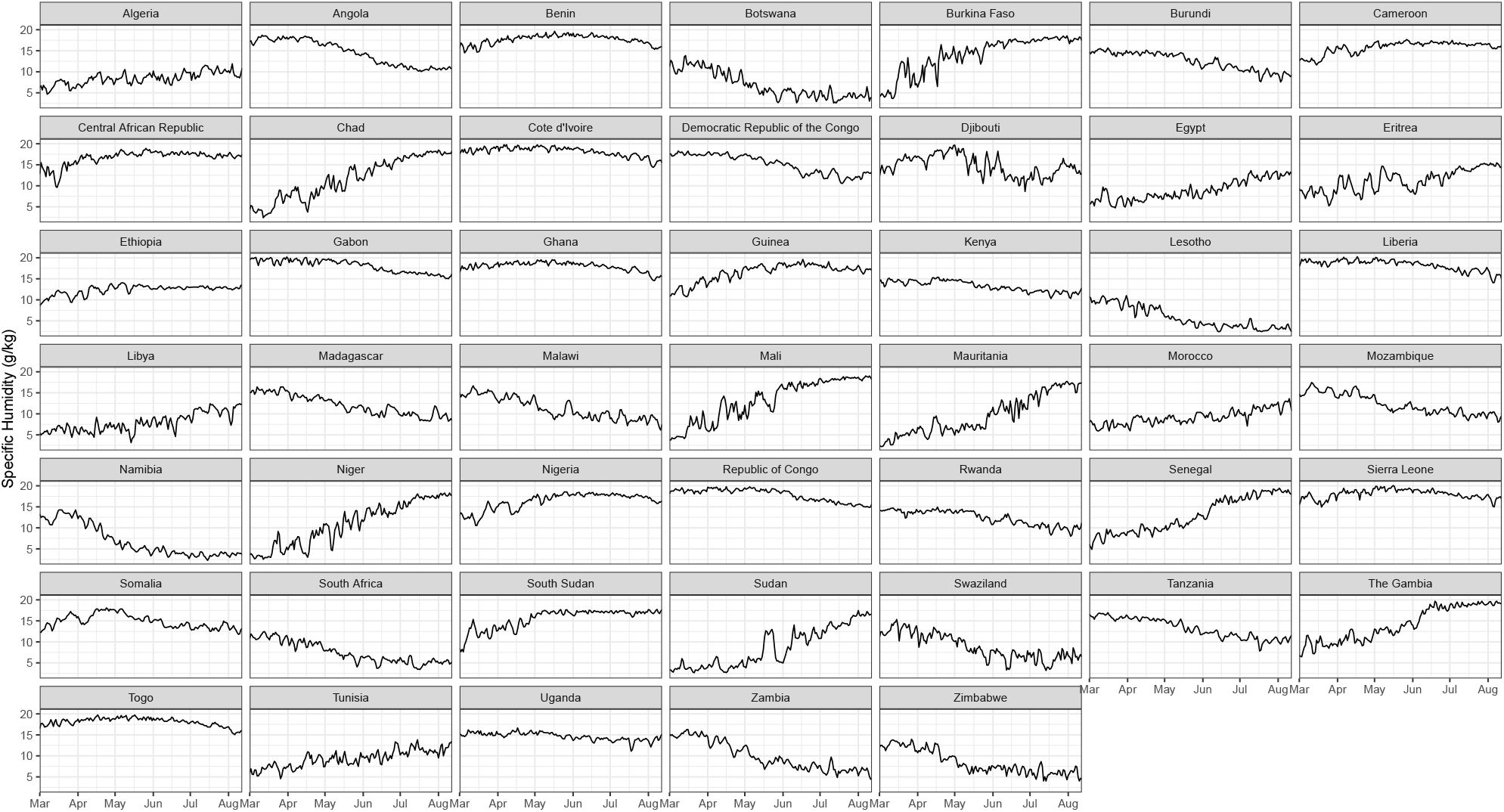
Time series of population-weighted mean specific humidity for all countries of Africa.

**Figure S7:**
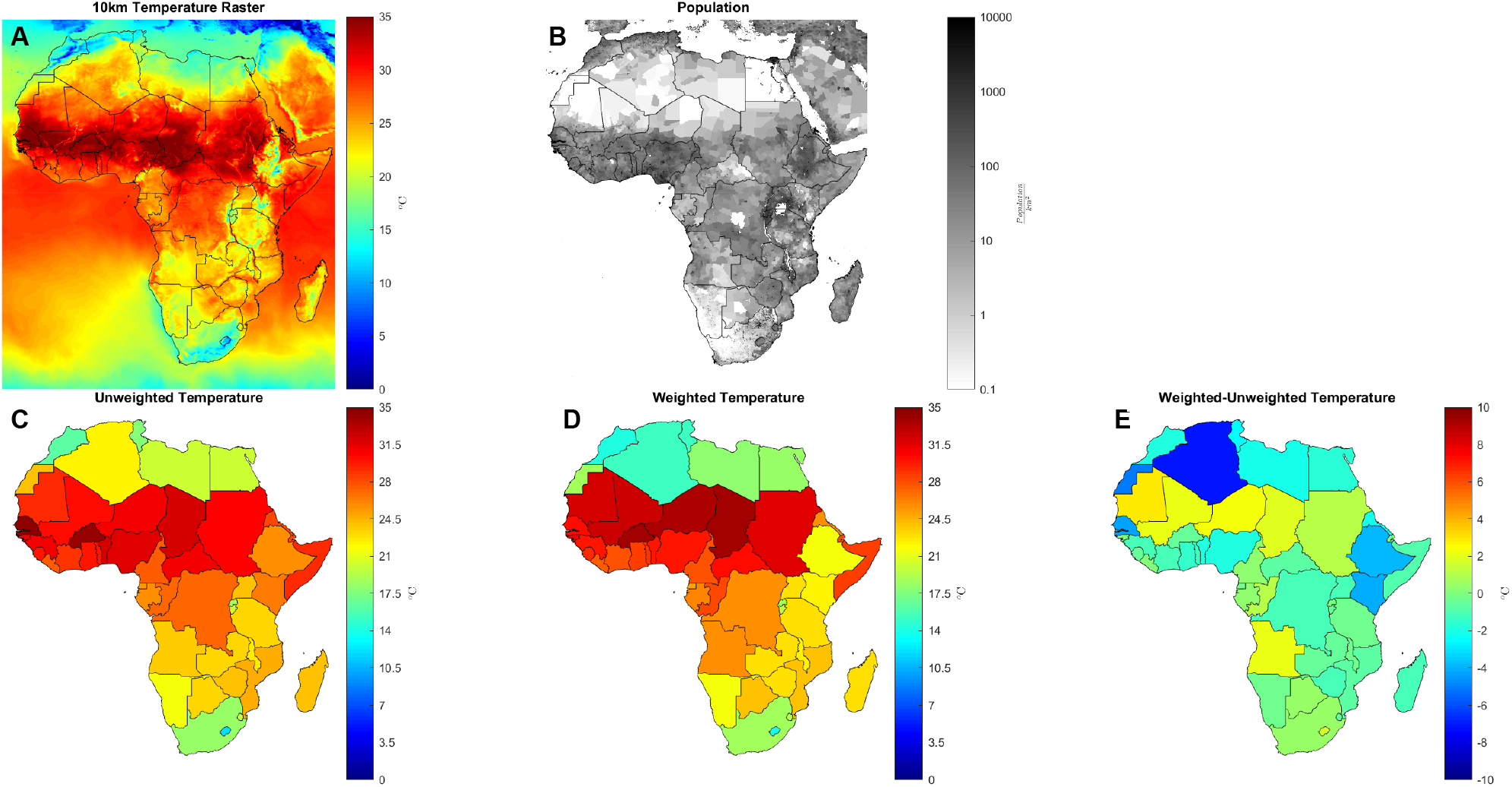
Population weighting of mean daily temperatures. (A) 10km mean temperature raster for April 1, 2020. (B) Gridded population density. (C) Unweighted mean temperature by country for the same day, taken as a simple average over each raster pixel within a country’s borders. (D) Mean temperature by country, weighted by population. (E) Differential of mean temperature by country between weighted and unweighted calculations.

**Figure S8:**
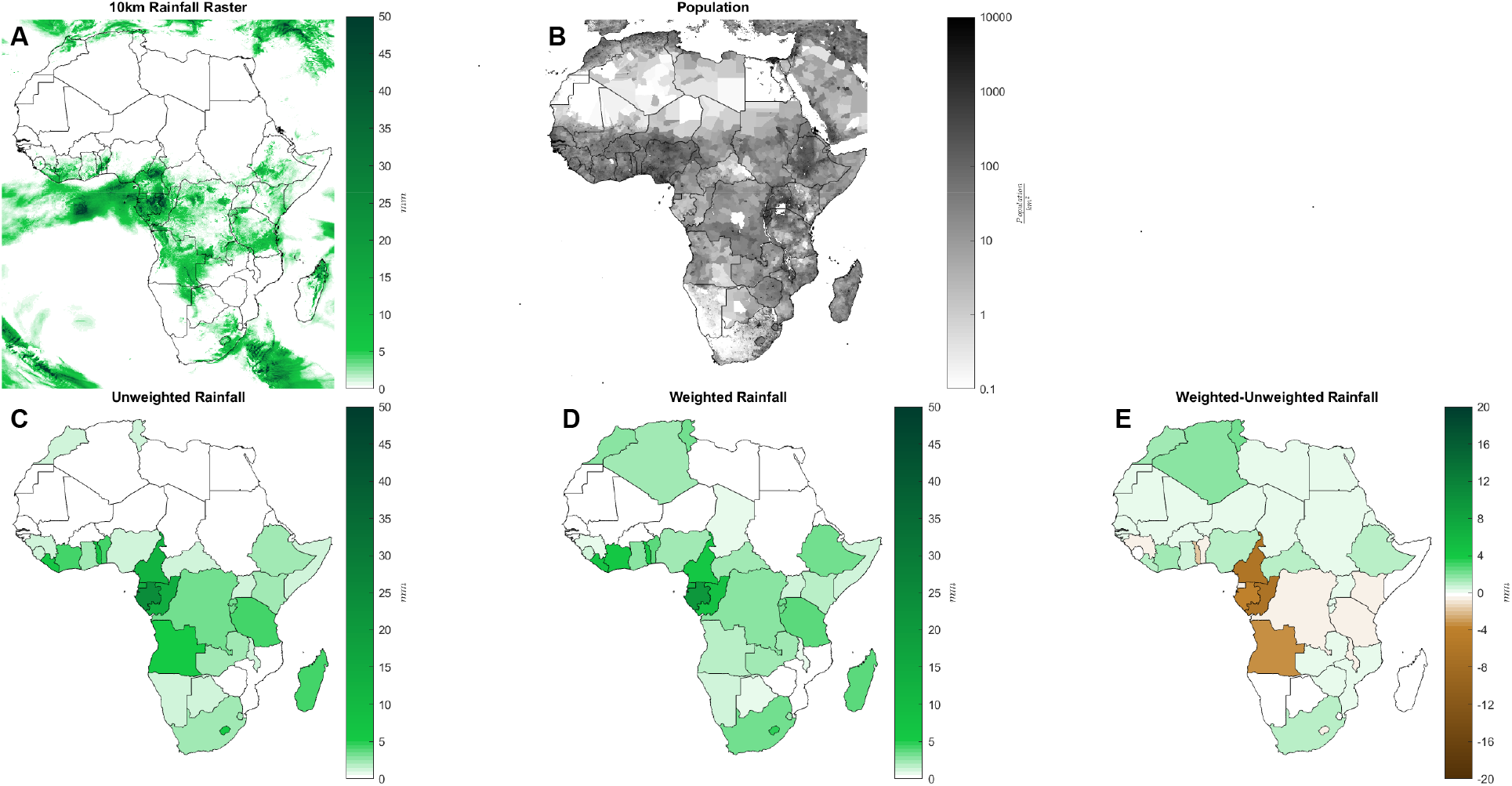
Population weighting of mean daily rainfall. (A) 10km mean rainfall raster for April 1, 2020. (B) Gridded population density. (C) Unweighted mean rainfall by country for the same day, taken as a simple average over each raster pixel within a country’s borders. (D) Mean rainfall by country, weighted by population. (E) Differential of mean rainfall by country between weighted and unweighted calculations.

**Figure S9:**
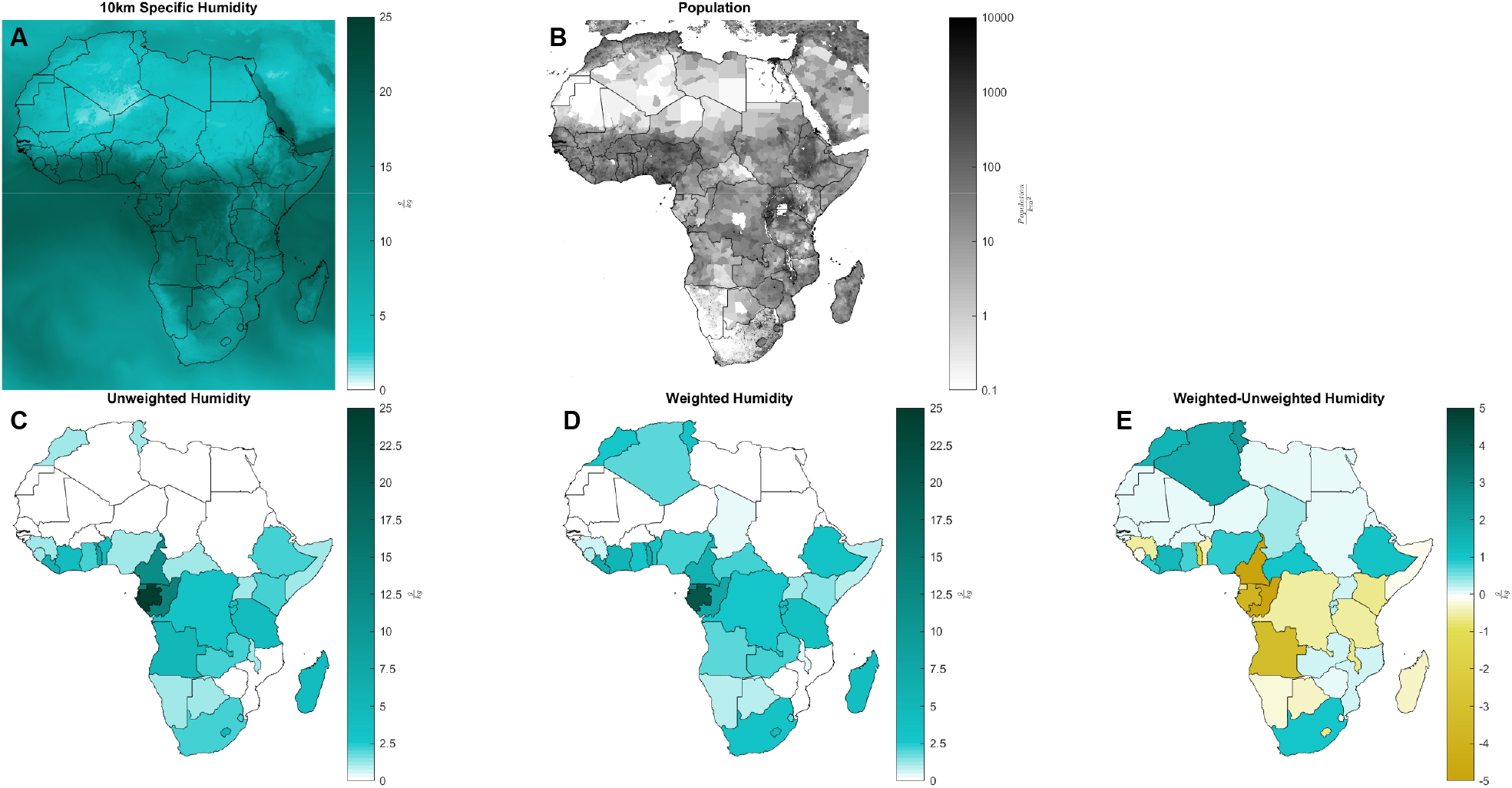
Population weighting of mean daily humidity. (A) 10km mean humidity raster for April 1, 2020. (B) Gridded population density. (C) Unweighted mean specific humidity by country for the same day, taken as a simple average over each raster pixel within a country’s borders. (D) Mean humidity by country, weighted by population. (E) Differential of mean specific humidity by country between weighted and unweighted calculations.

**Figure S10:**
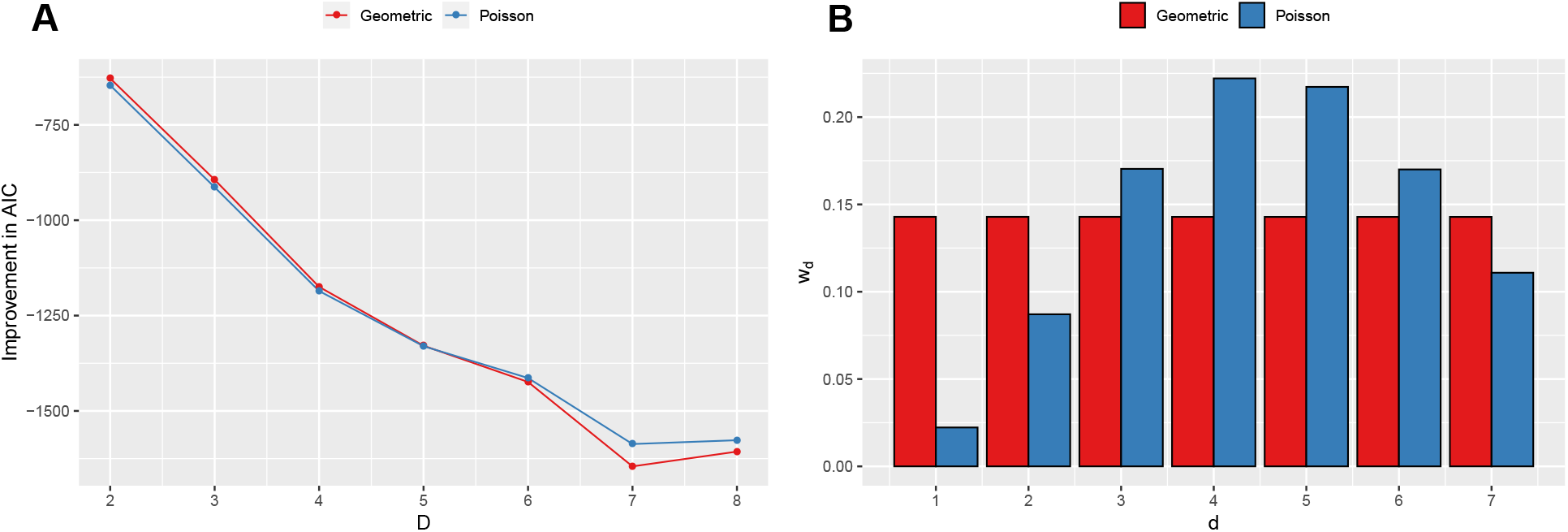
Distributed lags. (A) Improvement in AIC (relative to the first-order model, D = 1) for increasing lags and different weights parameterizations.The largest improvement (ΔAIC = -1560) is achieved by a model with t = 7 and with geometric weights. However we preferred the Poisson weights due to the weights distribution that is biologically consistent with the infection dynamics. AIC increases again at D = 8. For the final parsimonious model, we chose D=7 because of the largest improvement in AIC. (B) Estimated model weights 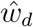. The Poisson weights show an increase in infectiousness up to day 4 followed by a decrease while geometric weights degenerate to a uniform distribution.

**Figure S11:**
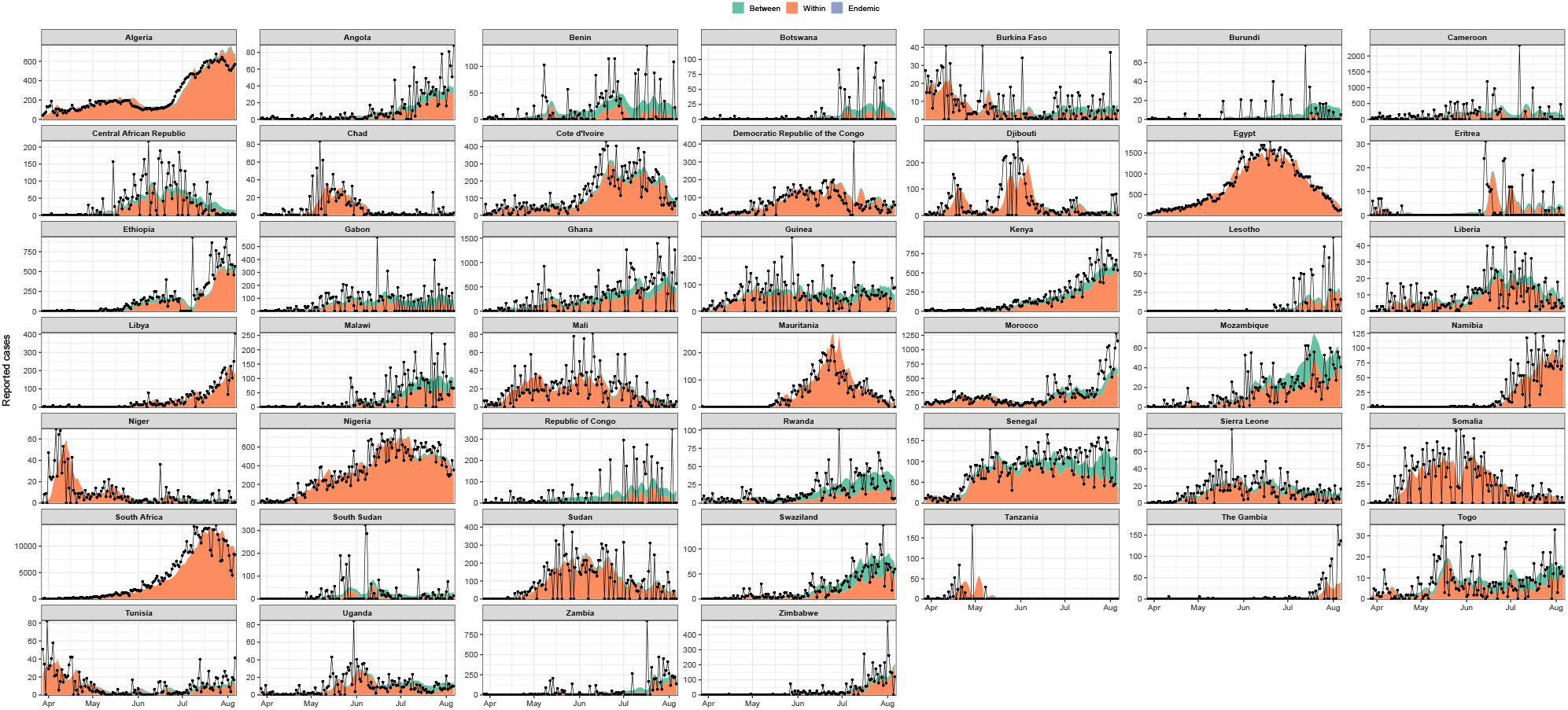
Model contributions by country. Observed case counts are shown with black traces. Between and within country components are shown for the period of analysis.

**Figure S12:**
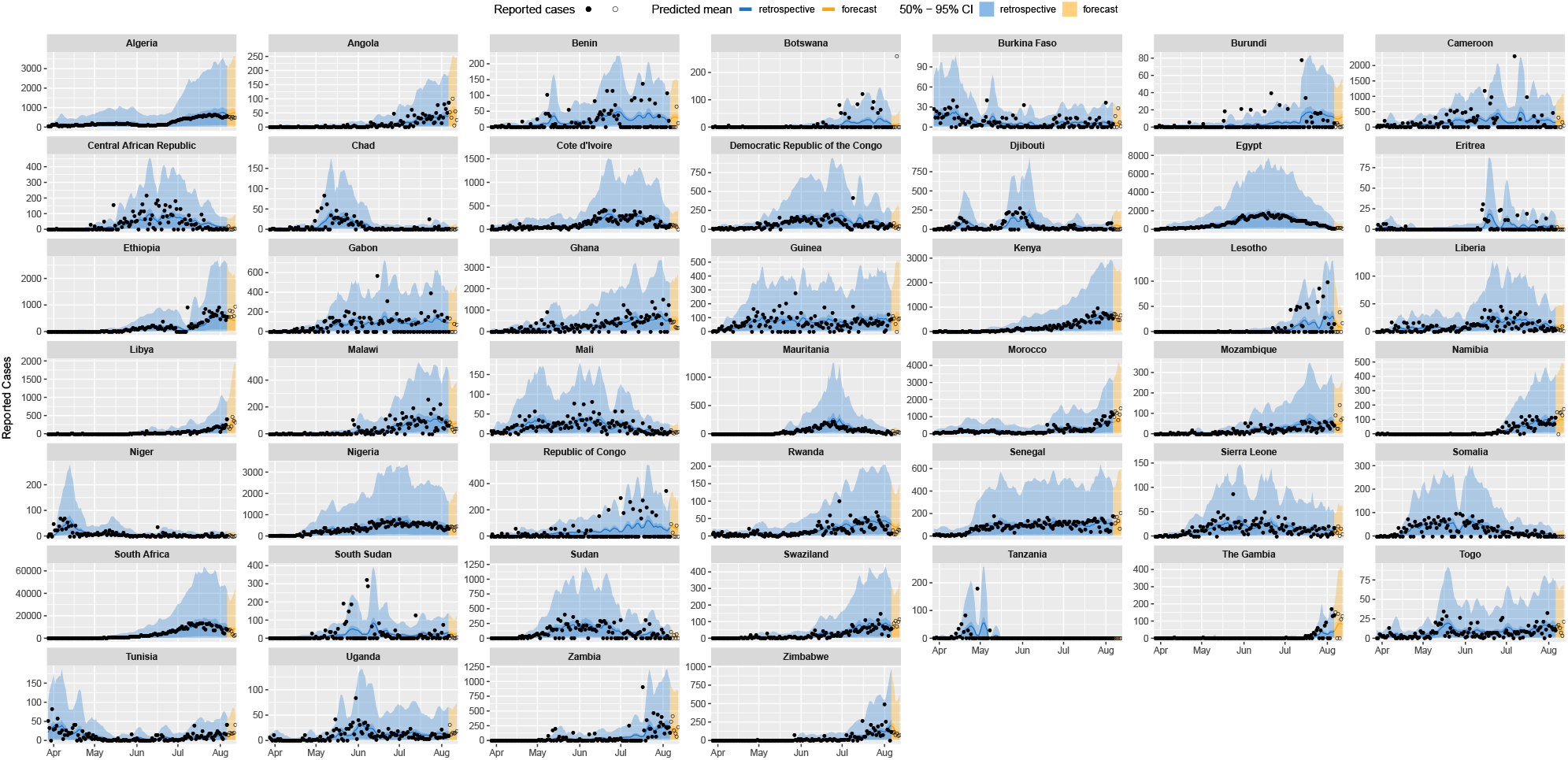
Week-ahead predictions by country. Reported cases used to fit the model are filled black circles. Unobserved cases from the period for which predictions are made are open black circles. The 50% and 95% confidence intervals are shown in dark/light shading for both the retrospective model fitting and the week-ahead forecast.

**Figure S13:**
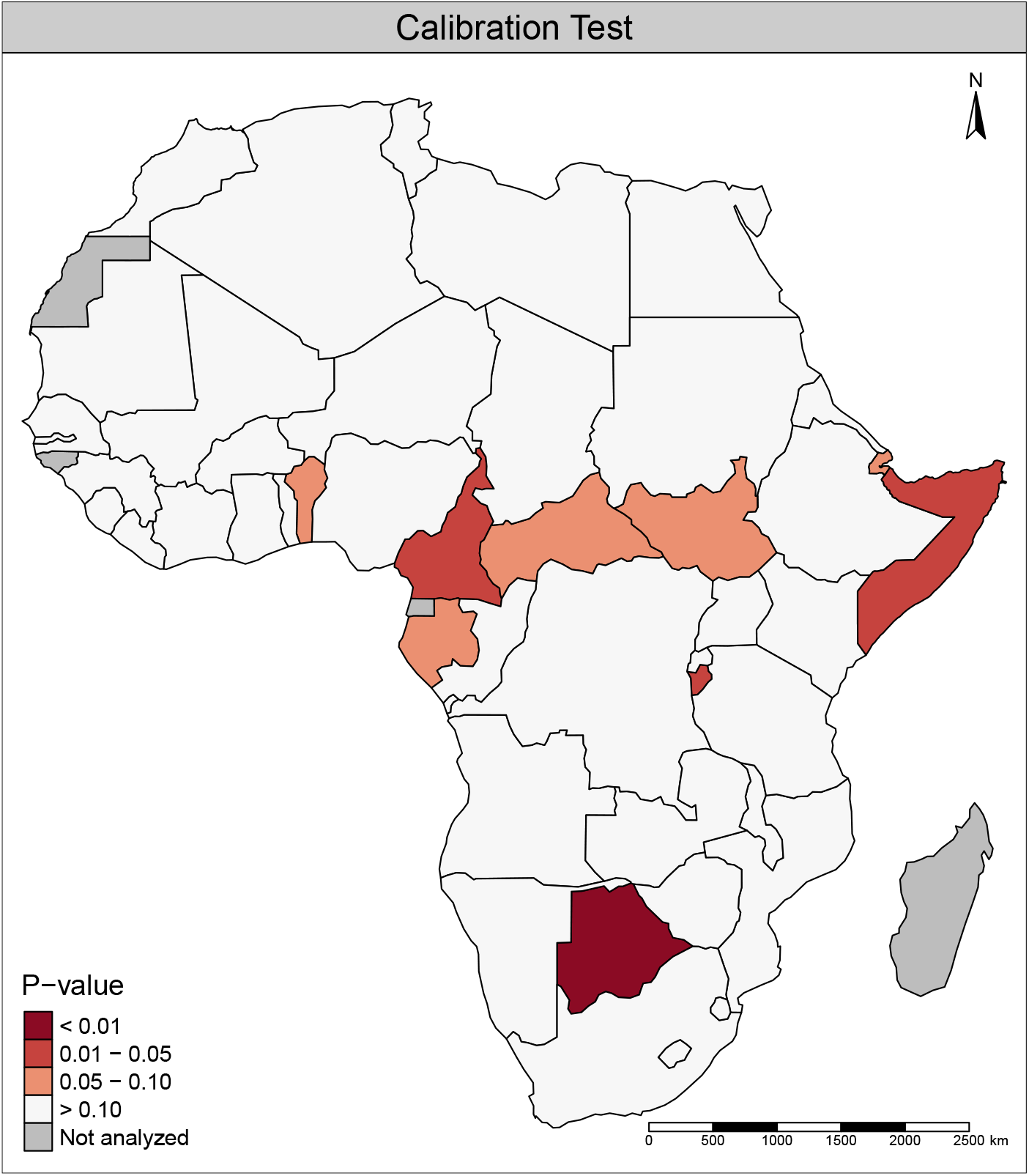
Calibration test results by country. P-value obtained from a calibration test performed on the hold-out data (last week of cases) for each country using the logarithmic score. Small p-values correspond to poorly calibrated predictions. A global test for the whole continent confirms that our model is well calibrated (*p* = 0.96).

**Figure S14:**
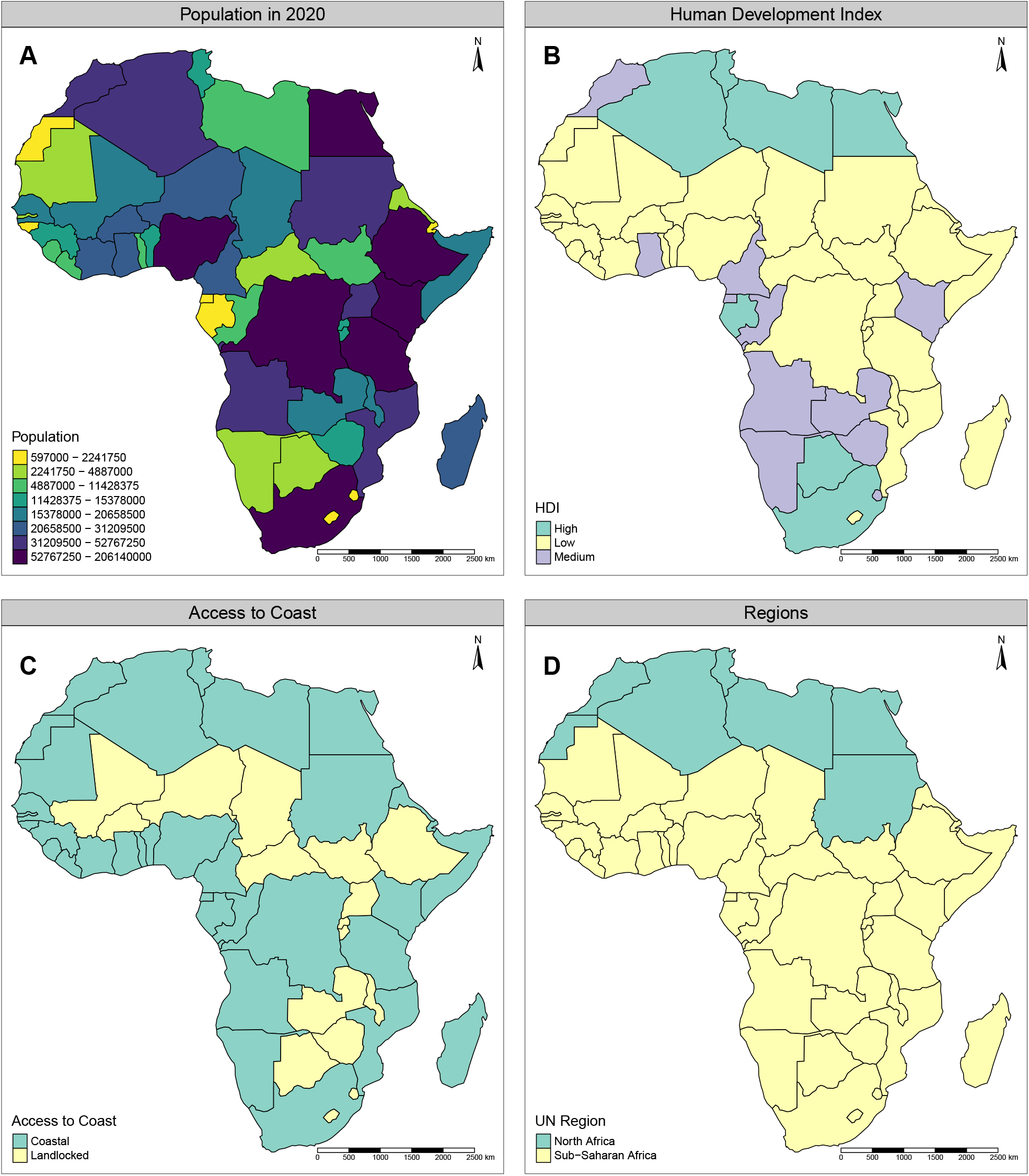
Time-constant covariates utilized in the model. (A) Population. (B) Human Development Index. (C) Access to coast. (D) North African and Sub-Saharan Africa regions.

**Figure S15:**
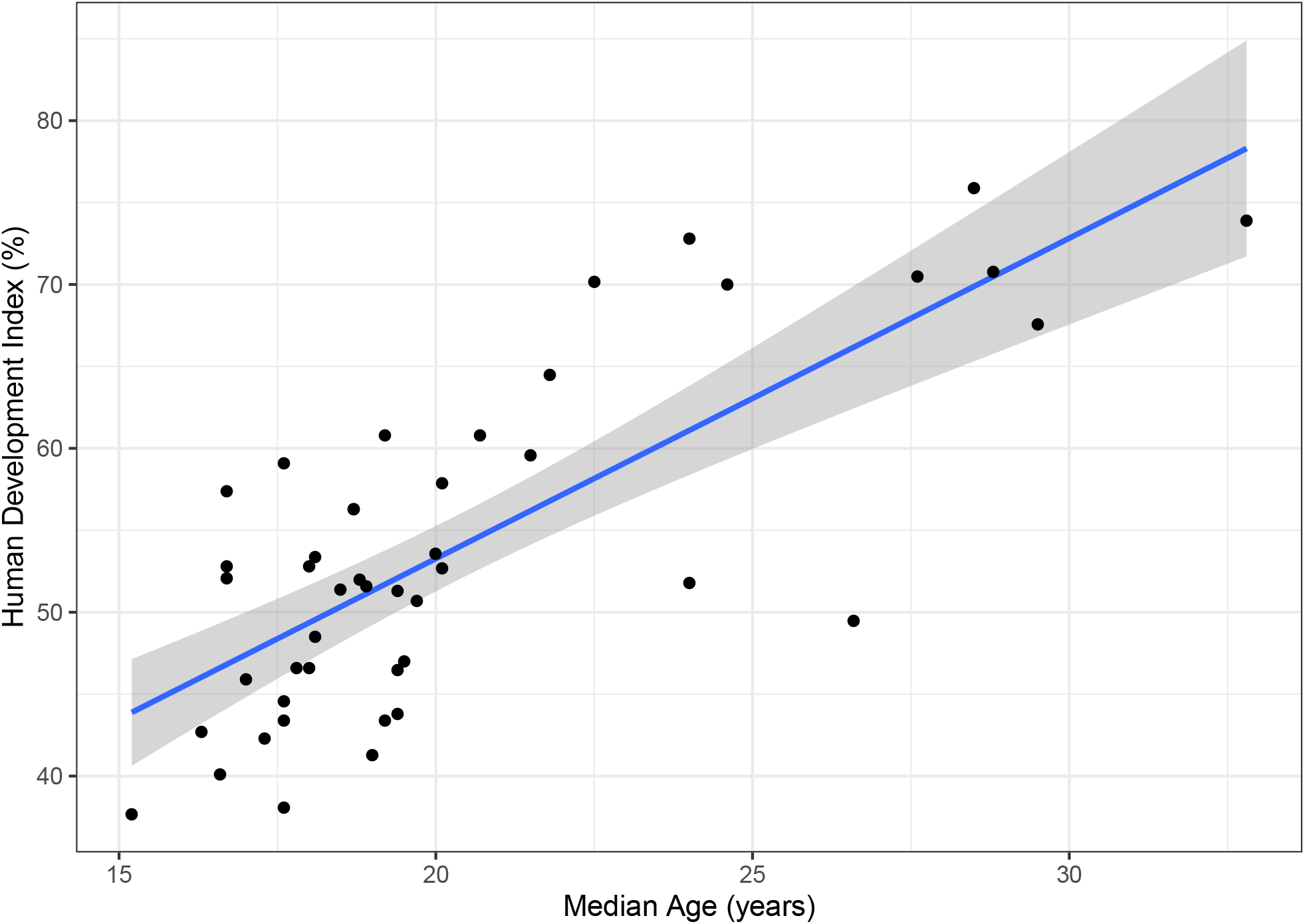
Correlation of human development index of a country with median age. The Pearson’s correlation coefficient is high (R=0.71, p*<*0.0001). To avoid multicolinearity with HDI, we removed median age from the model fit.

**Figure S16:**
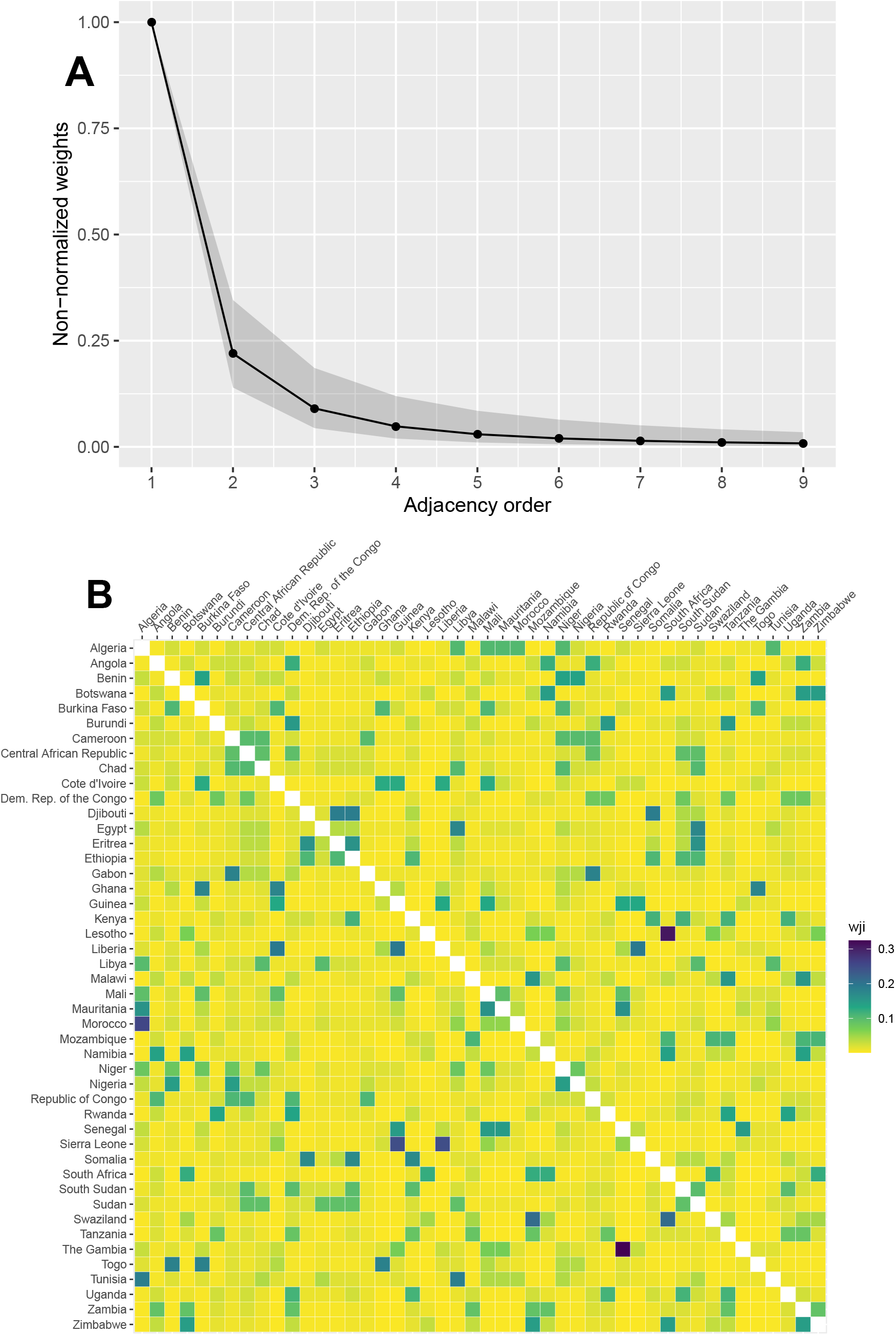
Spatial connectivity of countries. (A) Spatial connectivity weights as a function of country border distance. (B) A matrix of countries showing the connectivity weights. The connectivity weights decrease as the number of borders crossed to reach a certain country increase.

**Table S1:** Model selection for AR7: Sequential model building from intercept-only model (model 1) to the final model (model 5). Statistically significant explanatory factors are in **bold**.

| Variable | Model 1 |  | Model 2 |  | Model 3 |  | Model 4 |  | Model 5 fixed effects |  | Model 5 (Final) with random effects |  |
| --- | --- | --- | --- | --- | --- | --- | --- | --- | --- | --- | --- | --- |
|  | RR | 95 % CI | RR | 95 % CI | RR | 95 % CI | RR | 95 % CI | RR | 95 % CI | RR | 95 % CI |
| **Endemic** |  |  |  |  |  |  |  |  |  |  |  |  |
| Intercept | **33.707** | 21.473 - 52.91 | **11.83** | 4.12 - 33.972 | **16.33** | 7.416 - 35.959 | **23.371** | 13.275 - 41.144 | **27.949** | 19.407 - 40.252 | **11.071** | 7.15 - 17.142 |
| **Within-country** |  |  |  |  |  |  |  |  |  |  |  |  |
| Intercept | 0.988 | 0.939 - 1.039 | 0.93 | 0.783 - 1.104 | **0.513** | 0.374 - 0.705 | **0.521** | 0.38 - 0.714 | 0.767 | 0.552 - 1.064 | 0.958 | 0.05 - 1.669 |
| log(population) |  |  | 0.981 | 0.944 - 1.018 | 0.977 | 0.939 - 1.017 | 0.975 | 0.936 - 1.015 | 0.99 | 0.949 - 1.032 | 1.036 | 0.956 - 1.123 |
| HDI |  |  |  |  | 1.054 | 0.973 - 1.141 | 0.991 | 0.913 - 1.075 | 0.995 | 0.914 - 1.083 | 0.957 | 0.819 - 1.118 |
| Landlocked |  |  |  |  | **0.874** | 0.766 - 0.998 | **0.862** | 0.754 - 0.985 | **0.783** | 0.685 - 0.895 | **0.674** | 0.531 - 0.857 |
| Stringency |  |  |  |  | **1.91** | 1.379 - 2.645 | **1.94** | 1.405 - 2.679 | **1.674** | 1.195 - 2.345 | **1.872** | 1.170 - 3.00 |
| Testing |  |  |  |  | 1.058 | 0.994 - 1.127 | **1.066** | 1.002 - 1.134 | **0.925** | 0.863 - 0.991 | **0.817** | 0.729 - 0.918 |
| Rain |  |  |  |  | 1.033 | 0.977 - 1.093 | 1.034 | 0.978 - 1.092 | 1.019 | 0.963 - 1.078 | 1.045 | 0.981 - 1.112 |
| Temperature |  |  |  |  | **1.082** | 1.015 - 1.153 | **1.07** | 1.005 - 1.138 | 1.059 | 0.995 - 1.127 | **1.106** | 1.014 - 1.206 |
| Humidity |  |  |  |  | 0.99 | 0.925 - 1.06 | 0.984 | 0.92 - 1.052 | 0.982 | 0.918 - 1.051 | **0.856** | 0.780 - 0.940 |
| **Between-country** |  |  |  |  |  |  |  |  |  |  |  |  |
| Intercept | **0.027** | 0.024 - 0.03 | **0.424** | 0.267 - 0.673 | **0.435** | 0.274 - 0.692 | **0.34** | 0.207 - 0.561 | **0.025** | 0.014 - 0.046 | **0.045** | 0.04 - 0.481 |
| log(population) |  |  | **1.776** | 1.611 - 1.958 | **1.776** | 1.612 - 1.957 | **1.837** | 1.65 - 2.035 | **1.656** | 1.500 - 1.828 | 1.428 | 0.923 - 2.21 |
| HDI |  |  |  |  |  |  | **1.746** | 1.486 - 2.052 | **1.71** | 1.45 - 2.017 | 1.239 | 0.584 - 2.638 |
| Landlocked |  |  |  |  |  |  | 1.117 | 0.902 - 1.384 | **1.35** | 1.082 - 1.685 | 0.844 | 0.306 - 2.323 |
| Stringency |  |  |  |  |  |  |  |  | **2.622** | 1.544 - 4.45 | 1.630 | 0.560 - 4.749 |
| Testing |  |  |  |  |  |  |  |  | **2.899** | 2.482 - 3.385 | **2.322** | 1.676 - 3.218 |
| AIC | 48824.32 |  | 48690.89 |  | 48663 |  | 48618.73 |  | 48437.99 |  | - |  |
| $\rho$ | 0.64 | 0.43 - 0.85 | 0.79 | 0.55 - 1.04 | 0.86 | 0.62 - 1.10 | 0.64 | 0.38 - 0.89 | 0.74 | 0.45 - 1.02 | 2.19 | 1.53 - 2.84 |
| $\psi$ | 2.11 | 2.025 - 2.195 | 2.062 | 1.978 - 2.145 | 2.044 | 1.961 - 2.127 | 2.024 | 1.942 - 2.107 | 1.947 | 1.867 - 2.027 | 1.703 | 1.631 - 1.775 |
| Logarithmic Score | 4.772 |  | 4.817 |  | 4.809 |  | 4.765 |  | 4.745 |  | 4.713 |  |

**Table S2:**
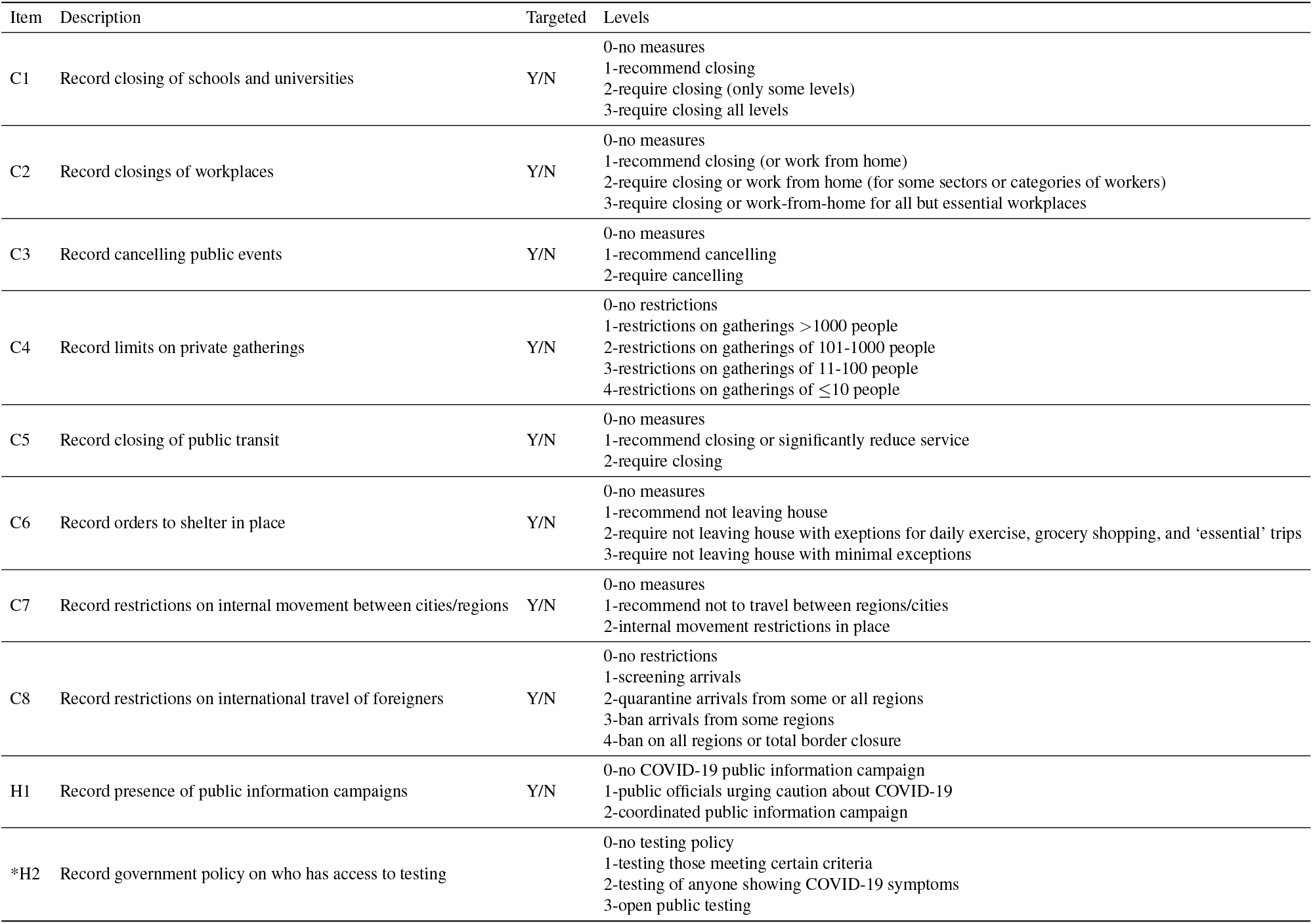
Codebook for the factors included in the OxCGRT stringency index. Full codebook [here](https://github.com/OxCGRT/covid-policy-tracker/blob/master/documentation/codebook.md), and methodology for stringency index calculation given [here](https://github.com/OxCGRT/covid-policy-tracker/blob/master/documentation/index_methodology.md). *Testing policy H2 does not contribute to the stringency index. It is included as a separate variable in the model.

**Table S3:** Sources for data included in the model. Results presented in this work can be reproduced with code and archived versions of this data that have been stored in a [github repository](https://github.com/SchiffLab/COVID19-HHH4-Africa). Current versions of this data can be found at the links included.

| **Data** | **Source** |
| --- | --- |
| Country-level COVID-19 incidence | Johns Hopkins University the Center for Systems Science and Engineering (JHU-CSSE) Coronavirus Resource Center — JHU-CSSE Website — Data Repository. |
| Country Population, 2020 | United Nations (UN) Population Division of the Department of Economic and Social Affairs — UN Website |
| Geography - Shapefiles | ESRI: Version November 21, 2018. This shapefile has been modified from the original ESRI version to represent the current UN country designations. This modified shapefile has been included in the data repository. |
| Geography - Sub-Saharan Africa | United Nations Statistics Division (UNSD) — UNSD Website |
| Geography - Landlocked | UNSD — UNSD Website |
| Government Containment Policies | Oxford COVID-19 Government Response Tracker (OxCGRT) — OxCGRT Website — Data Repository |
| Human Development Index | United Nations Development Programme (UNDP) Human Development Index for 2018 — UNDP Website & Data |
| Weather data | UK Met Office — UK Met Office Website see methods section for procedure to extract population-weighted weather variables |

## Notes

### Competing Interest Statement

The authors have declared no competing interest.

### Author Declarations

This study relied on publicly available case data and did not require IRB oversight.

